# Bridging Genomic Insight and Clinical Care in Chromosome 8p Disorders Through a Registry-Driven Passport

**DOI:** 10.64898/2025.12.02.25340619

**Authors:** Tobias Brünger, Kaiti Syverson, Bina Maniar, Jacob Borello, Scott Demarest, Lauren Chaby, Dennis Lal

## Abstract

**Background:** Rare diseases are frequently associated with prolonged diagnostic odysseys and fragmented care, requiring coordination across multiple specialties and often leaving families to bridge gaps in medical knowledge among providers. Chromosome 8p disorders, caused by diverse structural rearrangements, exemplify these challenges, as clinical manifestations and management needs vary widely across genetic subgroups. Although natural history studies (NHSs) systematically collect longitudinal and genotype-phenotype data, these insights are seldom translated into practical, patient-centered tools that directly inform day-to-day clinical care.

**Results:** We developed the 8p Patient Passport, an automated tool that generates individualized clinical and genomic summaries and anchors each finding within subgroup-specific reference data derived from the Project 8p NHS. Each passport integrates detailed genomic information, including precise 8p rearrangement coordinates, together with developmental, behavioral, and comorbidity data, and presents these findings in the context of the individual’s representative genetic subgroup. The automated R/LaTeX pipeline produces personalized, plain-language summaries for families and providers within one minute per case. Forty-two passports were generated across three molecular subgroups and distributed to the families that participated in the NHS: inverted duplication/deletion (n = 30), interstitial deletion (n = 10), and duplication (n = 2). In a post-deployment survey, caregivers rated the Passport highly for clarity (mean 4.8/5), accessibility (4.8/5), and usefulness (4.2/5), emphasizing its value for communicating with healthcare and educational teams and for understanding their child’s presentation in relation to peers.

**Conclusions:** The 8p Patient Passport bridges rare-disease research and clinical care by transforming NHS data into individualized, context-aware care tools. By pairing patient-level data with subgroup-specific reference distributions, it supports personalized interpretation, facilitates care coordination, and empowers families as active participants in medical decision-making. This framework lays the groundwork for scalable, interoperable applications across rare-disease communities, aligning with global efforts to advance patient-centered precision medicine.

## Background

Rare diseases comprise a heterogeneous group of more than 7,000 clinically distinct conditions that continue to expand with advances in genetic diagnostics. Collectively, they affect approximately 5-6% of the global population, yet each disorder occurs in only a small number of individuals, creating unique diagnostic and management challenges within healthcare systems^1^. Patients frequently endure prolonged “diagnostic odysseys” marked by years of uncertainty and repeated misdiagnoses, reflecting the limited availability of clinical expertise, insufficient access to comprehensive genetic testing, and the inherent complexity and heterogeneity of these conditions^2^. Even after a diagnosis is established, care often remains fragmented, requiring coordination among multiple specialists across disciplines, a task that commonly falls to families^3^. Emergency and frontline healthcare providers are typically unfamiliar with rare conditions, which can lead to suboptimal management during acute encounters. As a result, families frequently act as both medical educators and care coordinators, spend considerable time explaining essential medical information to unfamiliar care teams, and often struggle to communicate their specialized care needs while trying to integrate services across a fragmented healthcare landscape^4,5^.

Chromosome 8p disorders exemplify these broader challenges within rare disease management. These neurodevelopmental conditions arise from diverse structural genomic rearrangements affecting the short arm of chromosome 8, including inverted duplication-deletions, terminal or interstitial deletions, duplications, trisomies, translocations, and mosaic events. Affected individuals commonly present with global developmental delays, intellectual disability, epilepsy, structural brain anomalies, congenital heart defects, and other systemic health issues^6^. Although these rearrangements often produce overlapping clinical phenotypes, recent genotype-phenotype studies have revealed key distinctions among genetic sub-disorders. For example, chromosome 8p deletions involving the *GATA4* gene are significantly associated with congenital septal heart defects^7^. Such information is crucial for early surveillance and intervention but rarely communicated effectively to non-specialist clinicians, underscoring persistent challenges in translating genomic findings into clinical practice^8^. Consequently, two patients with apparently similar chromosome 8p rearrangements may differ markedly in clinical course, and without precise subgroup classification based on the underlying genomic architecture (e.g., deletion, duplication, or inverted duplication–deletion)^9^, individualized prognosis and management remain uncertain.

Natural history studies (NHSs) have become central to advancing rare disease research by systematically collecting standardized clinical, genetic, and patient-reported data to define genotype–phenotype correlations and disease trajectories^10–12^. Long-standing examples, such as lysosomal disease registries, have demonstrated how sustained, real-world data collection can inform monitoring guidelines and improve patient outcomes^13^. Despite their value, many NHSs struggle with participant enrollment and retention because families caring for high-needs children often experience substantial survey fatigue^14^. The burden of repeatedly completing lengthy online surveys can be especially high in NHSs that require longitudinal data collection. Providing a tangible benefit or immediate return for participating families, such as an individualized outputs or clinical summaries, can therefore meaningfully increase enrollment, retention, and satisfaction^15^. Recent initiatives aim to enhance the usefulness of registry data through advocacy-driven data portals, electronic health record integration, and patient-carried summaries that improve care coordination. The International Patient Summary (IPS) standard exemplifies this effort by providing a concise core dataset for cross-setting use, but it focuses on summarizing an individual’s medical information rather than placing it in the context of others with the same rare condition, and therefore does not provide disease-specific insights^16^.

In this context, we introduce the 8p Patient Passport, an automated tool that transforms NHS data into concise, individualized summaries provided directly to participants. Each passport integrates detailed genomic information, including precise 8p rearrangement breakpoints, with key clinical and developmental data summarized in plain language and contextualized against genetically defined subgroups. Automated generation through an R^17^/LaTeX^18^ pipeline reduces the workload for patient advocacy groups and enables rapid updates as patients age or new data are added. By presenting an accessible overview of clinical features and subgroup-specific risks, the passport empowers families to advocate effectively and supports clinicians in delivering informed and coordinated care. Although developed for chromosome 8p disorders, this framework is adaptable across other rare disease communities and translates complex registry data into practical, actionable tools for patient-centered care.

## Methods

### Study Design, Ethical Oversight, Participant Recruitment, and Data Management

The Chromosome 8p Natural History Study is an ongoing global study integrating data that is retrospective, prospective, caregiver-reported, and clinician-reported from families with chromosome 8p rearrangements. Its objectives are to characterize genotype–phenotype relationships, symptom trajectories, and multisystem involvement in chromosome 8p disorders to translate into precision care tools and targeted interventions for individuals affected by chromosome 8p disorders (IRB# NB200051). Participants are recruited internationally through the Project 8p Foundation network, clinician referrals, and community outreach. Families enrolled in the 8p NHS, together with their supporting providers, contributed to defining the Passport’s content and data priorities to ensure alignment with community needs. Informed consent is obtained from all participants or their legal guardians prior to enrollment. Data are collected through modular online surveys capturing demographic, clinical, developmental, treatment, and genetic information, complemented by uploaded medical records and assessments such as MRI and EEG reports. Participation is voluntary, and individuals can withdraw at any time without consequence. All data is stored on a secure, access-controlled, GDPR^19^- and HIPAA^20^-compliant registry platform using end-to-end encryption during transmission and at rest. Each participant is assigned a unique de-identified identifier to facilitate downstream analyses and 8p Patient Passport generation. Access to identifiable information is restricted to authorized personnel under least-privilege access policies. Routine data integrity checks, including completeness assessments, logical-consistency reviews, and duplicate detection, are performed before harmonization and analysis to ensure that only de-identified, quality-controlled datasets are included in subsequent processing and reporting.

### Automated Data Processing and Passport Generation

The 8p Patient Passport pipeline converts raw NHS exports from multiple survey modules into harmonized, de-identified participant summaries, which are then used to generate individualized PDF passports through a parameterized R (v4.4.0)^17^ Markdown and LaTeX^18^ framework. During harmonization, all survey variables were mapped to a unified schema, synonymous items were merged, and categorical responses were standardized to consistent value labels (for example, “yes,” “Yes,” or “1” were consolidated as “present”). This modular harmonization framework is adaptable to other rare disease registries, as variable mappings and reference tables can be readily extended or customized for additional genes or disorder-specific cohorts. Genetic information was similarly normalized: reported chromosome 8p copy-number events were aligned to GRCh38 genomic coordinates, start and end positions were standardized, and cytobands were assigned using reference boundaries. Each participant’s genetic subgroup, such as an inverted duplication with terminal deletion, was inferred through deterministic logic based on reported intervals and survey responses.

### 8P Patient Passport Issuance and Satisfaction Survey

Following the distribution of the 8p Patient Passport, a post-deployment satisfaction survey was conducted to assess usability, accessibility, and perceived clinical value among families who participated in the 8p NHS and had received an 8p Patient Passport. The survey included multiple-choice and open-ended questions addressing ease of access, clarity and comprehensibility, usefulness for families and healthcare providers, perceived value of individual sections, suggestions for improvement, and likelihood of future use in clinical or educational settings. Responses were collected anonymously through the registry platform and summarized descriptively. All questions of the survey are accessible in the Supplementary Table 1. Feedback and user suggestions are being incorporated into upcoming Passport versions to guide iterative improvement.

## Results

### Generation of the 8p Patient Passport

We developed the 8p Patient Passport, an automated, individualized clinical report designed to support caregivers and clinicians in improving care and quality of life for individuals with chromosome 8p rearrangements (referred to within the Project 8p community as “8p Heroes”). Each 8p Patient Passport begins with a one-page *Key Information* summary that outlines the patient’s genetic diagnosis, major risk factors, provider guidance, and recommended next steps, providing an at-a-glance overview of essential findings. Subsequent sections integrate patient-specific chromosome 8p breakpoint coordinates, developmental milestone trajectories, neurobehavioral and comorbidity profiles, medication and supplement histories, and prenatal and birth information collected through the 8p NHS. All data are contextualized against subgroup-specific reference distributions to provide actionable clinical insights.

Passports are generated directly from the 8p NHS, a patient advocacy–driven study using standardized and custom surveys that capture array- or sequencing-derived 8p copy-number variant (CNV) breakpoints together with comprehensive phenotypic data, including developmental milestones, clinical symptoms, therapies, and medication use (see Methods). Each report is produced automatically in less than one minute through an R and LaTeX pipeline, yielding a median document length of eight pages (Figure 1). In total, 42 individuals were issued 8p Patient Passports comprising the reference cohort. Participants were classified into three molecular subgroups: inverted duplication/deletion (n = 30), duplication (n = 2), and interstitial deletion (n = 10). Example Passports for each subgroup (8p deletion; 8p duplication and 8p inverted deletion duplication) are provided in the Supplementary data file.

**Figure 1.**
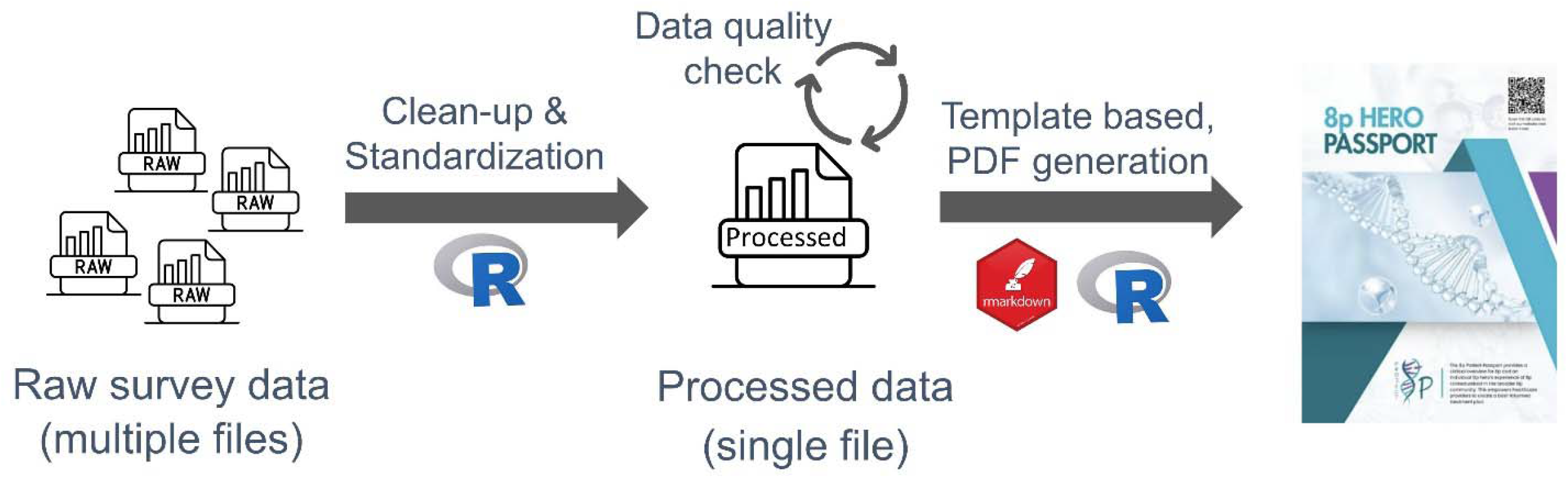
Workflow for generating a personalized 8p Patient Passport.

### Genomic, Developmental, and Clinical Panels of the 8p Patient Passport

Each 8p Patient Passport is organized into three core panels: *Genomic Diagnosis, Developmental and Therapeutic Overview*, and *Clinical Symptom Profile*. Together, these panels provide a unified summary of an individual’s genomic architecture, developmental trajectory, treatment history, and multisystem involvement relative to peers within the same genetic subgroup.

The *Genomic Diagnosis* panel visualizes the precise 8p CNV breakpoints of each individual in the context of their genetic subgroup, allowing direct comparison with the distribution of rearrangements observed across the cohort. This view highlights both the individual’s unique genomic CNV and its position within the broader spectrum of 8p rearrangements. For example, among individuals with inverted duplication/deletion (invdupdel) CNVs, deletions ranged from 6 to 58 Mb (median 24.2 Mb) and duplications from 2 to 12 Mb (median 6.8 Mb), demonstrating considerable structural variability even within a single subgroup (Figure 2). Such visual context enables clinicians to recognize whether a patient’s genomic profile lies within or outside the typical range for their genetic subgroup, providing a framework for discussing genotype-phenotype associations and clinical expectations.

**Figure 2.**
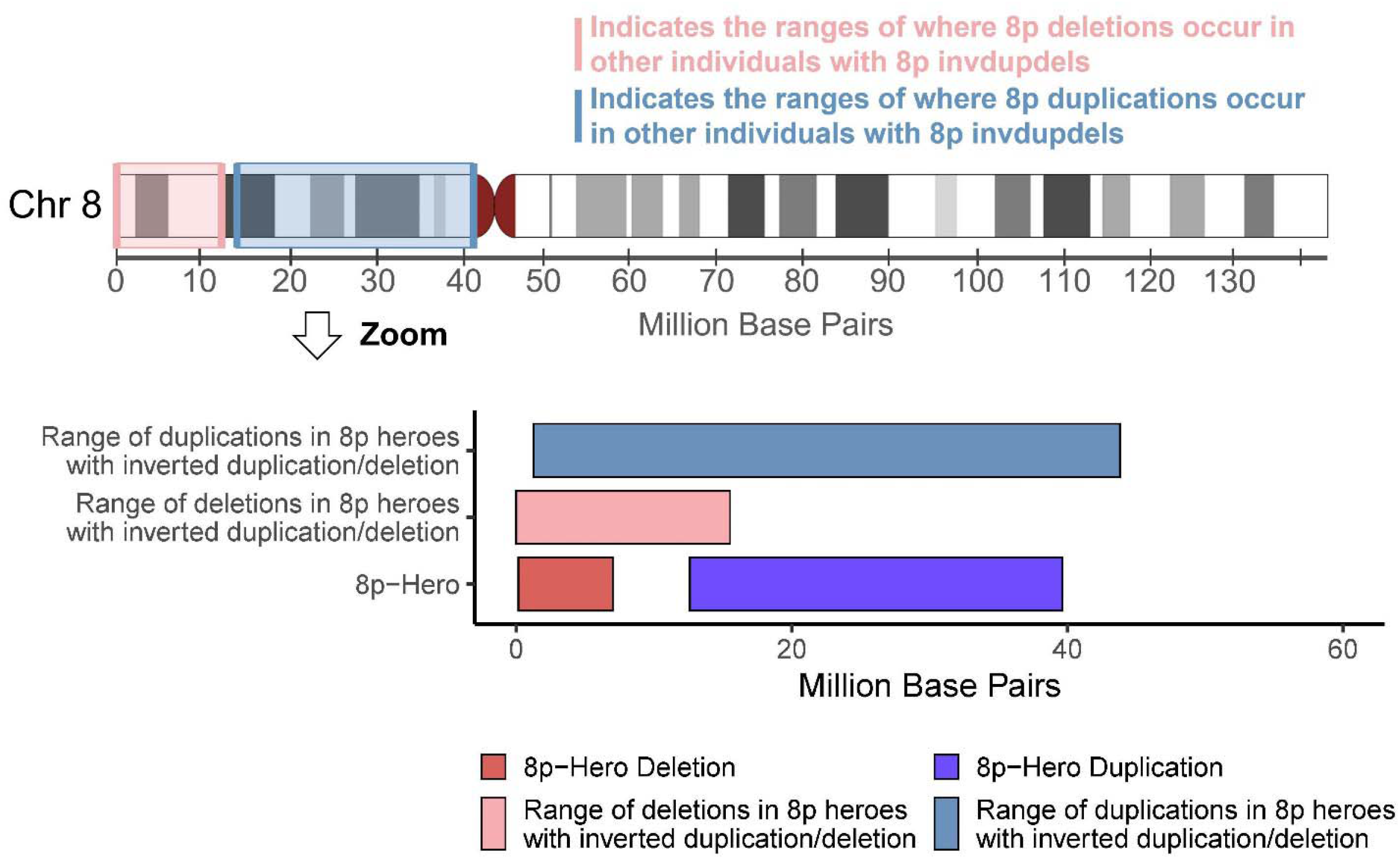
Genomic ranges of deletions and duplications in 8p Heroes with inverted duplication/deletions (invdupdel). Distribution of deletions and duplications along chromosome 8p in participants with invdupdel. The upper schematic shows the short arm of chromosome 8 (hg38) with shaded regions indicating the positions where deletions (pink) and duplications (blue) occur across the cohort. The lower bar plots compare the ranges and median sizes of these rearrangements, illustrating deletions spanning 6-58 Mb (median 24.2 Mb) and duplications spanning 2-12 Mb (median 6.8 Mb). The index case is highlighted in red and blue, representing a 27 Mb deletion and a 6.8 Mb duplication.

The *Clinical Symptom Profile* panel of the 8p Patient Passport integrates developmental, physical, and neurobehavioral data to provide a concise overview of an individual’s clinical presentation within their genetic subgroup. Key milestones, including first syllables, first words, sentence formation, crawling, and walking, are compared with subgroup-specific percentile distributions (Figure 3A). In the example 8p Patient Passport, several milestones were achieved later than the subgroup median, indicating developmental delay consistent with the broader neurodevelopmental impact of chromosome 8p rearrangements. The Clinical Symptom Profile further summarizes co-occurring manifestations across domains. Within the genetic subgroup, symptoms such as hypotonia, motor coordination difficulties, and growth or vision abnormalities (Figure 3B) are commonly observed. The made-up individual of the example 8p Patient Passport displays a pattern consistent with the reference group. Similarly, neurobehavioral characteristics including intellectual disability, repetitive behaviors, and reduced social initiation are prevalent among individuals with an invdupdel and are likewise represented in the illustrated case (Figure 3D). Comparing each individual’s profile with subgroup-level frequencies highlights both expected and atypical features, supporting clinicians in recognizing comorbidities and anticipating care needs. Medication information (Figure 3C) as part of the *Developmental and Therapeutic Overview* complements this profile by listing the individual’s current treatments, which in this example include anti-seizure agents such as valproic acid and felbamate together with supportive supplements such as melatonin and vitamin D. Integrating developmental, clinical, and therapeutic information supports individualized care planning and facilitates communication across specialties.

**Figure 3.**
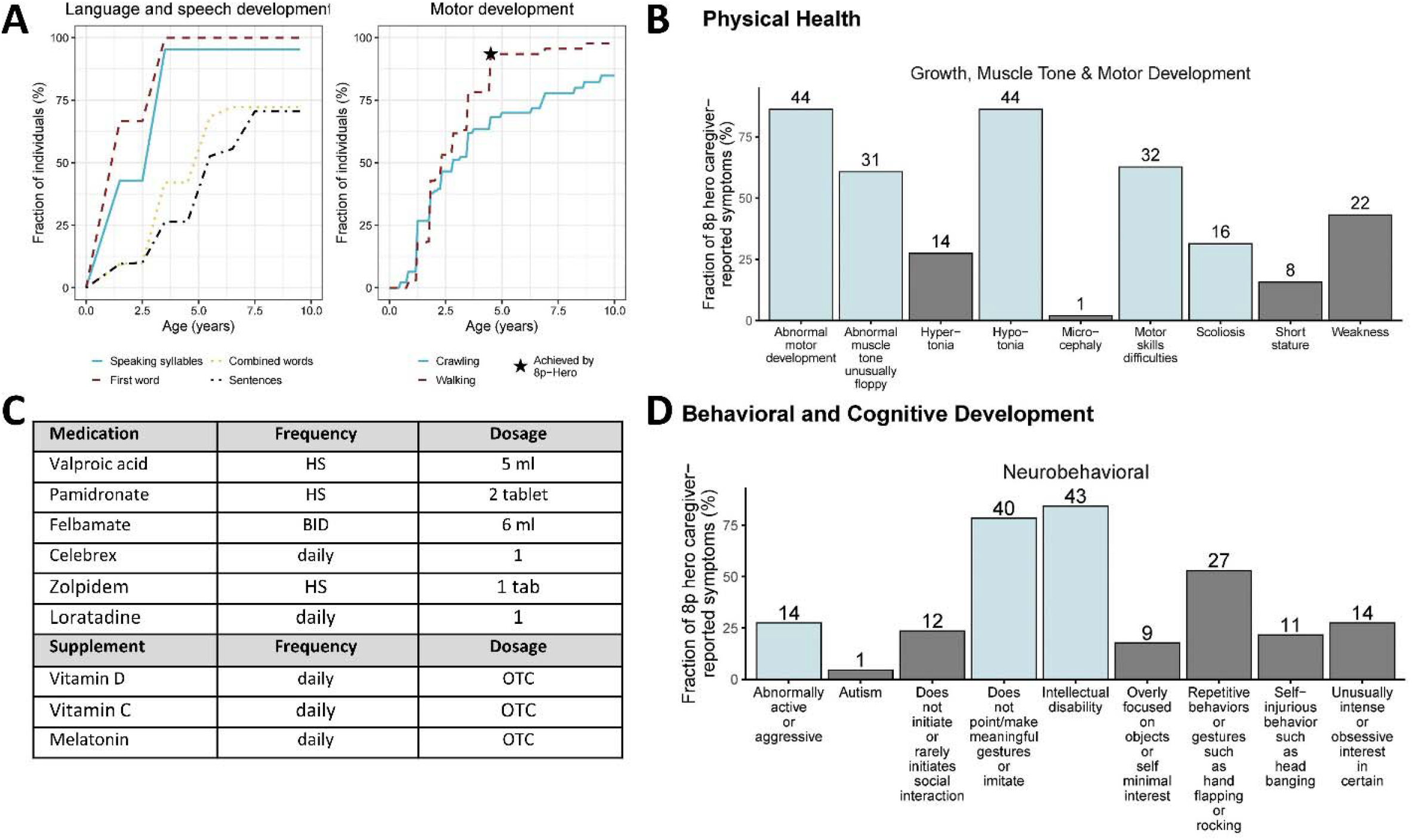
Developmental, physical health, and neurobehavioral features in 8p Heroes. This figure illustrates how the Clinical Symptom Profile component of the 8p Patient Passport summarizes an individual’s developmental and clinical presentation in comparison with peers from the same genetic subgroup. (A) Developmental milestones, including early language and motor skills, are displayed relative to subgroup-specific percentile distributions, with the illustrated individual showing delayed attainment across several domains. (B) Physical findings such as hypotonia, coordination difficulties, and growth or vision abnormalities are common within the subgroup and are also observed in the example individual. (C) Current medications are listed, including anti-seizure agents (valproic acid, felbamate) and supportive supplements (melatonin, vitamin D). (D) Neurobehavioral characteristics like intellectual disability, repetitive behaviors, and reduced social initiation are frequent among individuals with an inverted duplication/deletion (invdupdel) and are likewise represented in the illustrated case.

### User Feedback and Perceived Utility

A post-deployment survey of five families (12% response rate) demonstrated strong endorsement of the 8p Patient Passport as a useful and accessible care tool. On a five-point Likert scale, respondents rated the Passport as easy to access (mean 4.8), easy to understand (4.8), and helpful to families (4.2), with a high likelihood of continued use in clinical or educational settings (4.8; Figure 4). Participants reported that the Passport effectively reflected their 8p Hero’s experiences (mean 4.0) and highlighted the value of visualizing their child’s clinical features in relation to others with similar 8p rearrangements. When asked which sections were most valuable, all respondents selected the Genomic Summary and How to Connect with My 8p Hero sections, and four of five selected the Developmental Milestones, Physical Health, and Behavioral and Cognitive Development summaries (Supplementary Figure 1). Open-ended comments highlighted the Passport’s clarity, its usefulness for communicating with healthcare and support professionals, and its value in illustrating how a child’s presentation relate to other individuals within the same genetic subgroup (see Supplementary Table 2 for full testimonials). Open-ended question feedback and suggestions from this survey will inform refinements in upcoming Passport versions.

**Figure 4.**
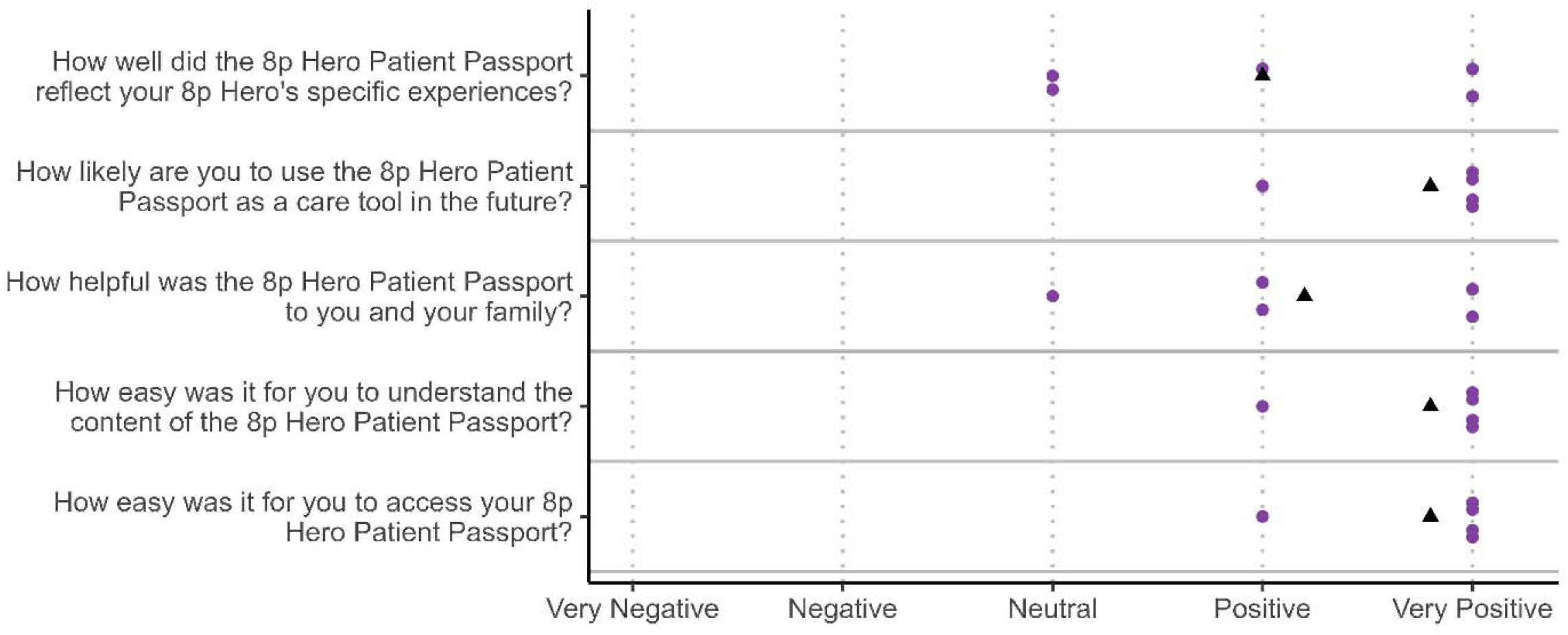
Caregiver ratings of the 8p Patient Passport. Responses from five families (12% response rate) evaluating the 8p Patient Passport on a five-point Likert scale ranging from Very Negative (1) to Very Positive (5). Each purple dot represents an individual response, and black triangles indicate the mean rating for each question. Items assessed included reflection of the participant’s experiences, ease of access, clarity of content, helpfulness to families, and likelihood of future use. Higher scores denote greater perceived accessibility, clarity, and usefulness of the Passport as a care tool.

## Discussion

The 8p Patient Passport transforms data from the 8p NHS into individualized clinical summaries that bridge rare-disease research and everyday care, using chromosome 8p disorders as a model for broader rare-disease applications. Feedback from families participating in the 8p post-deployment survey underscores the Passport’s utility in (i) facilitating communication with healthcare providers, educators, and caregivers, (ii) improving their understanding of complex 8p disorders consistent with prior work showing that structured patient-carried summaries are perceived as valuable tools for navigating fragmented care landscapes^5^, and (iii) providing an immediate return for their time investment in NHS participation. Each 8p Patient Passport compiles individualized genomic, developmental, and treatment information paired with cohort-derived benchmarks, creating a portable, disease-specific reference that supports communication across care settings. The 8p Patient Passport demonstrates how a rare disease NHS can be transformed into an actionable point-of-care tool, aligning with global calls for more personalized, patient-centered care^21,22^.

Rare disease management is often fragmented, as individuals typically rely on multiple specialists and support systems. Many providers encounter only a few patients with any specific rare condition during their careers, leaving families to bridge critical knowledge gaps. For chromosome 8p disorders, this has historically meant that caregivers bear the responsibility of educating clinicians about the condition. In one survey, more than 95% of rare disease caregivers reported spending substantial time explaining their child’s medical needs to new providers, and two-thirds found these interactions challenging^5^. The 8p Patient Passport directly addresses this gap by providing a concise, validated summary of an individual’s condition and care needs. This tool mitigates provider unfamiliarity and ensures that essential information, including the genetic diagnosis, comorbidities, and emergency considerations, is readily accessible. Families often describe the repeated need to explain their child’s complex condition as frustrating and emotionally exhausting, reflecting the strain of assuming an expert role in clinical interactions^23^.

By providing a trusted, standardized summary, the 8p Patient Passport reframes these encounters: caregivers become informed partners, and clinicians can align their assessments with disease-specific norms, fostering more efficient, confident, and collaborative interactions.

Large reference cohorts are essential for advancing rare-disease care by providing the contextual data needed to interpret individual presentations. Registry-based and NHS consolidate heterogeneous clinical and molecular information, allowing risk stratification, early outcome prediction, and refinement of management guidelines^24,25^. For clinicians, these reference distributions transform isolated findings into actionable insights by showing how a patient’s development, comorbidities, or genomic features align with, or diverge from, the reference cohort presentation. However, the clinical utility of these datasets depends on their ability to capture intra-disease heterogeneity and define subgroups with distinct trajectories and care needs. In chromosome 8p disorders, recent genotype-phenotype studies have shown that subtle structural differences, such as inverted duplication/deletions versus interstitial deletions, correlate with distinct neurodevelopmental and systemic profiles^9^. Recognizing such subgroup-level variation is critical for translating findings from the NHS into individualized, precision-oriented care, ultimately informing targeted surveillance, therapy planning, and prognosis. The 8p Patient Passport operationalizes this principle by pairing patient-specific genomic and phenotypic data with subgroup-specific reference distributions, thereby providing clinicians with an interpretable framework to guide care decisions. Although developed for chromosome 8p disorders, this framework is broadly transferable to other rare-disease registries aiming to deliver patient-centered, data-driven decision-support tools.

While the 8p Patient Passport demonstrates clear benefits, several limitations warrant consideration. First, the content provided in the 8p Patient Passport depends on ongoing participation in the 8p NHS, which may result in incomplete or outdated information if families discontinue engagement. However, providing families with a personalized output may strengthen long-term participation and encourage timely data updates, an advantage over traditional registries that offer limited feedback and provide little immediate value to participants. Second, the tool currently relies primarily on caregiver-reported data, which can introduce biases or misclassification. For example, caregivers may conflate related medical terms, such as types of visual impairment. To mitigate this, the surveys of the NHS were written in accessible language, and participants received guidance materials explaining how to interpret and report clinical and genetic information. Unlike earlier rare-disease passport efforts that depend on free-text patient input and manual updating^5^, this framework addresses key limitations by providing an automated, reproducible pipeline linked to harmonized registry data, enabling rapid regeneration as new information becomes available. Future versions of the 8p Patient Passport aim to incorporate electronic health record integration and clinician-reported data, enabling automated updates and improved clinical validation while preserving usability and scalability.

## Conclusion

The 8p Patient Passport introduces a new model for rare-disease care by transforming NHS data into practical, patient- and clinician-facing tools. Using harmonized data from the 8p Natural History Study and an automated, reproducible pipeline, it generates individualized summaries that empower families in clinical communication and support informed decision-making. This model demonstrates how natural-history studies can deliver immediate clinical value while establishing a foundation for precision medicine and therapeutic development. Its structured data, GRCh38-aligned coordinates and modular pipeline establish a framework for future interoperability with standardized formats such as the HL7 FHIR International Patient Summary^26^ and emerging rare-disease common data models^27^. Ultimately, the 8p Patient Passport exemplifies personalized, data-informed medicine that bridges research and clinical practice and highlights the potential for registry-based frameworks to evolve into practical, scalable decision-support tools.

## Data Availability

Example 8p Patient Passports for all major genetic subgroups of individuals with 8p rearrangements are provided in the Supplementary data. The code and R/LaTeX pipeline used to generate the Passports is available at https://github.com/TobiasBruenger/8p-Patient-Passport/ alongside with randomly samples input files to exemplify the application of the code. Access to the underlying data from the Project 8p Natural History Study can be requested from the Project 8p Foundation (contact) and is subject to data use agreements and ethical approvals.

## List of abbreviations

NHS: Natural History Study;
CNV: Copy-Number-Variants;
invdupdel: inverted duplication/deletions

## Declarations

### Ethics approval and consent to participate

The Project 8p Natural History Study was approved by the Institutional Review Board of Nationwide Children’s Hospital (IRB# NB200051). All participants, or their legal guardians in the case of minors, provided informed consent prior to enrollment.

### Consent for publication

Consent for publication was obtained from all participants or their legal guardians as part of enrollment in the Project 8p Natural History Study, which includes permission to publish deidentified clinical, developmental, and genomic information. All data presented in this manuscript are fully de-identified.

### Availability of data and materials

Example 8p Patient Passports for all major genetic subgroups of individuals with 8p rearrangements are provided in the Supplementary data. The code and R/LaTeX pipeline used to generate the Passports is available at https://github.com/TobiasBruenger/8p-Patient-Passport/ alongside with randomly samples input files to exemplify the application of the code.

Access to the underlying data from the Project 8p Natural History Study can be requested from the Project 8p Foundation (contact) and is subject to data use agreements and ethical approvals.

### Competing interests

The authors report no conflicts of interest.

## Acknowledgement

We are deeply grateful to all families who participated in the Project 8p Natural History Study and generously shared their time and experiences. We also thank the caregivers, clinicians, and community members who contributed data and feedback through the surveys, helping to shape and improve the 8p Patient Passport. Their commitment and collaboration were essential to the success of this project.

## Funding

T.B. received a Project 8p Postdoctoral Fellowship.

## Authors’ contributions

Conceptualization: T.B., L.S., D.L.; Data curation: T.B., J.B, K.S; Analysis: T.B; Supervision: L.S, D.L., B.M.; Writing-original draft: T.B., L.S., D.L.; Writing-editing: D.L., L.S., K.S, J.B., B.M., S.D.

## Notes

### Competing Interest Statement

The authors have declared no competing interest.

### Author Declarations

The Institutional Review Board of Nationwide Children's Hospital gave ethical approval for this work. The Project 8p Natural History Study was reviewed and approved under IRB number NB200051, and all participants or their legal guardians provided informed consent prior to enrollment. All data analyzed for the 8p Patient Passport manuscript were de-identified prior to processing and reporting.

