## Supplementary data (Figure S1; Table S1-S2) for "Bridging Genomic Insight and Clinical Care in Chromosome 8p Disorders Through a Registry-Driven Passport"

**Supplementary Table 1.** Full questionnaire for the 8p Patient Passport issuance and satisfaction survey

| Question Text | Response Type |
|---|---|
| Timestamp | Automatically recorded |
| Role in community | Multiple choice |
| Could you provide a testimonial about the Patient Passport that Project 8p can use for awareness and improvements? | Open-ended |
| How easy was it for you to access your 8p Hero Patient Passport? | 5-point Likert |
| Do you have any comments on your ability to access your 8p Hero Patient Passport? | Open-ended |
| How easy was it for you to understand the content of the 8p Hero Patient Passport? | 5-point Likert |
| Do you have any feedback or suggestions about the clarity of the content? | Open-ended |
| How helpful was the 8p Hero Patient Passport to you and your family? | 5-point Likert |
| Do you have any comments on how helpful the Passport was or how it could be improved? | Open-ended |
| Which sections of the 8p Hero Patient Passport did you find the most useful? (Select all that apply) | Multiple choice |
| Did you find any part of the Passport unclear or confusing? | Yes/No |
| If yes, which parts? | Open-ended |
| How well did the 8p Hero Patient Passport reflect your 8p Hero's specific experiences? | 5-point Likert |
| Can you share more about how the 8p Hero Patient Passport did or did not reflect your 8p Hero's specific experiences? Are there any details you feel could better represent your Hero’s journey? | Open-ended |
| Have you shown the 8p Hero Patient Passport to one of your clinical providers, or do you plan to? | Open-ended |
| If yes, which providers have you shared it with or plan to share it with? | Open-ended |
| Did your provider(s) find the 8p Hero Patient Passport helpful or offer any feedback? | Open-ended |
| Were there any sections of the 8p Hero Patient Passport that you felt could be expanded or improved? | Yes/No |
| Which sections do you feel could be expanded or improved, and what specific changes would make them more helpful? | Open-ended |
| Are there any sections you would like to see added to the 8p Hero Patient Passport in the future? | Open-ended |
| How likely are you to use the 8p Hero Patient Passport as a care tool in the future? | 5-point Likert |

**Supplementary Table 2. Participant testimonials illustrating experiences with the 8p Patient Passport.** This section presents de-identified testimonials from families who completed the post-deployment satisfaction survey. Each statement reflects personal experiences with the 8p Patient Passport.

| |
|---|
| "It is a great initiative where other people can learn so much about your 8p hero. It is a tool that can be passed onto various professionals to help build a better picture of the child. I could clearly see how my chromosome disorder differs from that of others." |
| "The passport is a useful tool to share with medical professionals as well as therapists and caregivers." |
| "The patient passport is extremely helpful. I passed it on to my son's school team at his annual IEP meeting and they were very impressed." |
| "The Patient Passport is an excellent summary of my child for providers to quickly understand her symptoms in comparison to others with her genetic diagnosis. The passport has translated my participation in research into a useful tool to inform my child's providers and caregivers. This will contribute to an appreciation of my child." |

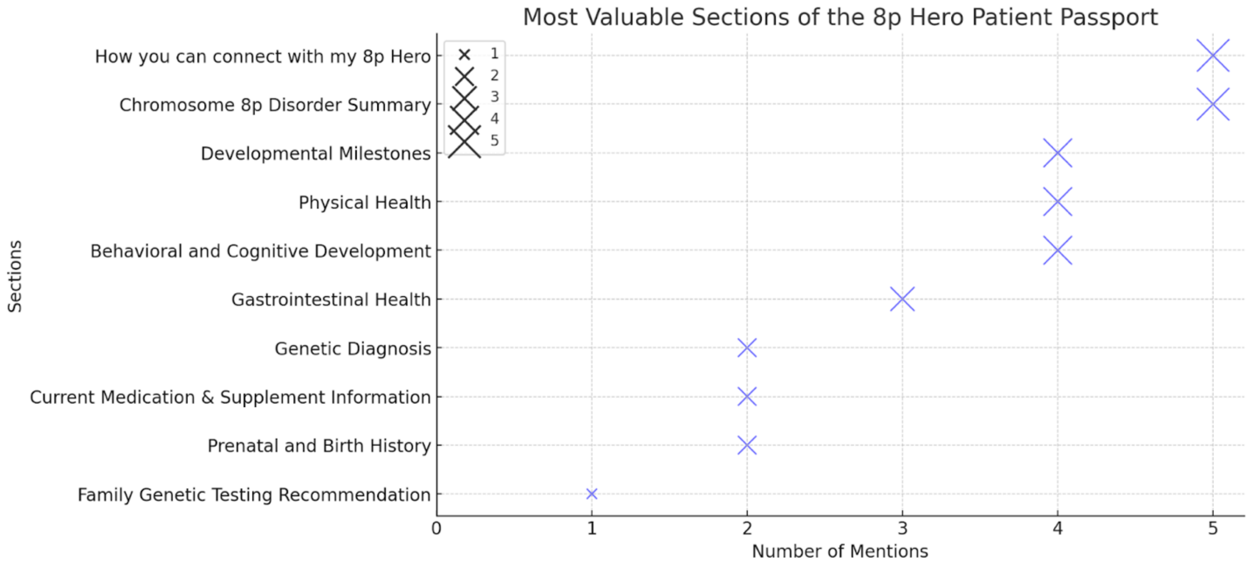


**Supplementary Figure 1. Most Valuable Sections of the 8p Hero Patient Passport.** This figure presents the most frequently identified valuable sections of the 8p Patient Passport, as reported by caregivers and individuals affected by Chromosome 8p rearrangements. Each point represents the number of respondents who found a given section useful. The size of each point corresponds to the relative frequency of selection. The x-axis displays the number of mentions, ensuring whole-number representation for clarity.

PROJECT 8P

### Example 8p Hero Passport: InvDupDel

**Name:**

**Date of birth:**

**Sex: Male** **Preferred Pronouns: Unknown**

#### Genetic Diagnosis

Date of testing: Feb-2020 Age of genetic testing: Unknown

Test type: Chromosomal microarray

Genetic finding: Chromosome 8p inverted duplication/deletion (8p deletion: 8p23.3 (156,048) to 8p23.1 (6,700,234); 8p duplication: 8p23.1 (11,604,632) to 8p11.22 (38,345,503))

Deletion size: 6544186 BP. Median size among other 8p heroes: 6,837,807 BP.

Duplication size: 26740871 BP. Median size among other 8p heroes: 24,194,174 BP.

Inheritance: Parental testing not done

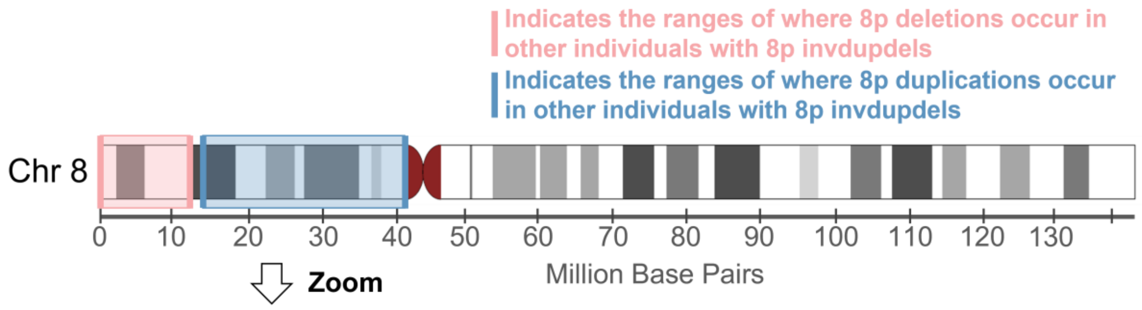


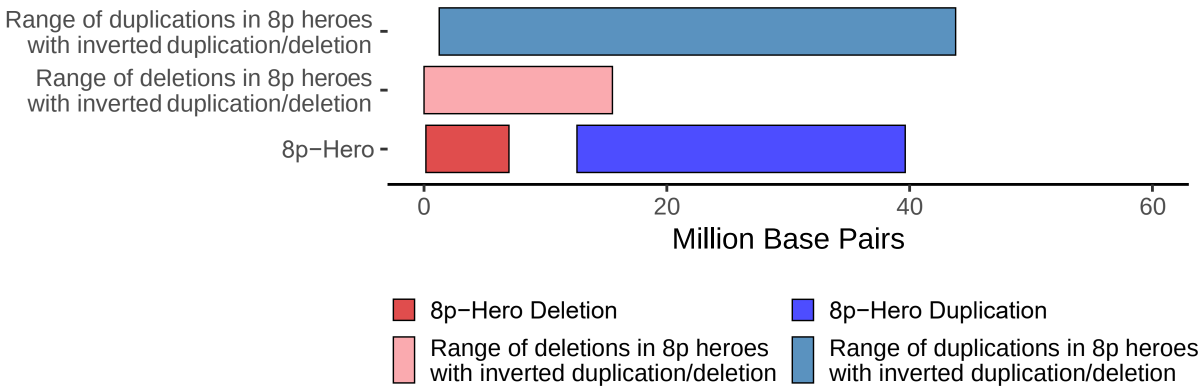


If you have any questions about any of the information provided in this 8p Hero Passport, or would like any of your information updated, please contact us at

#### Family Genetic Testing Recommendation

Given the genetic nature of chromosome 8p disorders, we recommend that family members consider genetic testing to better understand potential hereditary patterns. Family genetic testing can provide valuable insights into the unique genetic makeup of each family member, helping clinicians tailor care more effectively and enabling families to make informed decisions about health and wellness.

For more information on family genetic testing or to discuss options with a genetic counselor, please reach out to.

#### How you can connect with my 8p Hero

When interacting with my 8p Hero, it's important to approach with warmth, patience, and respect. Here are a few ways you can create a meaningful connection:

1. **Start with a greeting:** Say hello clearly and make eye contact when possible. Using my Hero's name helps them recognize your friendly intent.
2. **Pause for a response:** After asking a question or giving an instruction, wait for at least 3 seconds. My Hero may need extra time to process and respond, which could be in the form of a gesture, a nod, or even an eye blink.
3. **Use open body language:** Keep your gestures expressive and welcoming. A warm demeanor often encourages my Hero to feel comfortable in your presence.
4. **Respect personal space but be open to connection:** If my Hero shows interest in a hug or physical touch, depending on your comfort level, it's okay to offer a hand or a gentle hug.
5. **Other ways to connect to my 8p hero:**

#### Chromosome 8p Disorder Summary

Chromosome 8p disorder is a rare genetic condition that is known to affect 550 individuals worldwide, but has a reported prevalence of 1:10,000 - 1:30,000. Chromosome 8p disorder is caused when there is a rearrangement of genetic information before birth, on the short arm (the p arm) of the 8th chromosome. This genetic rearrangement can be the deletion of information, duplication of information, or inverted duplication and deletion (invdupdel). Chromosome 8p disorder has systemic effects, meaning it impacts cells and tissues throughout the body, rather than being confined to a single organ. The chromosomal rearrangement typically arises from spontaneous (de novo) changes early during embryonic development, for reasons that remain unclear. The severity and specific features of 8p disorders can vary significantly based on the amount of genetic information that has changed in addition to other factors that we are still working to understand.

There are several types of therapies that have been found to be effective for children with 8p disorders, depending on their symptoms, these include speech therapy, occupational therapy, physical therapy, vision therapy, ABA therapy, aquatic therapy, feeding therapy, hippotherapy as well as assistive and augmentative communication devices. In the absence of any treatments for chromosome 8p disorders, clinicians at the leading 8p multi-disciplinary clinic recommend treating symptoms with standard clinical practice as they arise.

**Currently, no targeted therapies exist for chromosome 8p disorders. As such, treatment focuses on managing symptoms as they appear, employing standard therapeutic interventions.** If you are treating a patient with chromosome 8p disorder and would like guidance or additional clinical insight from specialists in 8p, please reach out to our team. We can facilitate collaboration with our clinical experts to support your care plan and offer tailored resources. For further assistance, contact our clinical team at.

#### Current Medication & Supplement Information

| Medication | Frequency | Dosage |
| --- | --- | --- |
| Valproic acid | HS | 8 ml |
| Pamidronate | HS | 2 tablet |
| Felbamate | BID | 8 ml |
| Celebrex | daily | 1 |
| **Supplement** | **Frequency** | **Dosage** |
| Vitamin D | daily | OTC |
| Acidophilus | daily | OTC |

*Prior medications can be obtained from the Appendix.*

#### Prenatal and Birth History

| General information | Your 8p Hero | Other 8p Heroes with 8p inverted duplication/deletion median (25-75% quantiles) |
|---|---|---|
| Weight | 2500-4000g (5.5-8.8lbs) | - |
| Length | 49 cm 19.2 inch | 49cm (47cm - 50.8cm) 19.3 inch (18.5 inch - 20 inch) |
| Head Circumference | 34 cm 13.4 inch | 34cm (32cm - 35cm) 13.4 inch (12.6 inch - 13.8 inch) |
| Gestational | 37 - 40 weeks | - |
| **Prenatal & birth complications** | | |
| NICU Admission | No | 54.2% |
| NICU Reason | Unknown | - |
| Days at the Hospital following NICU Admission | - | 5 (3 - 16) |

#### Developmental Milestones

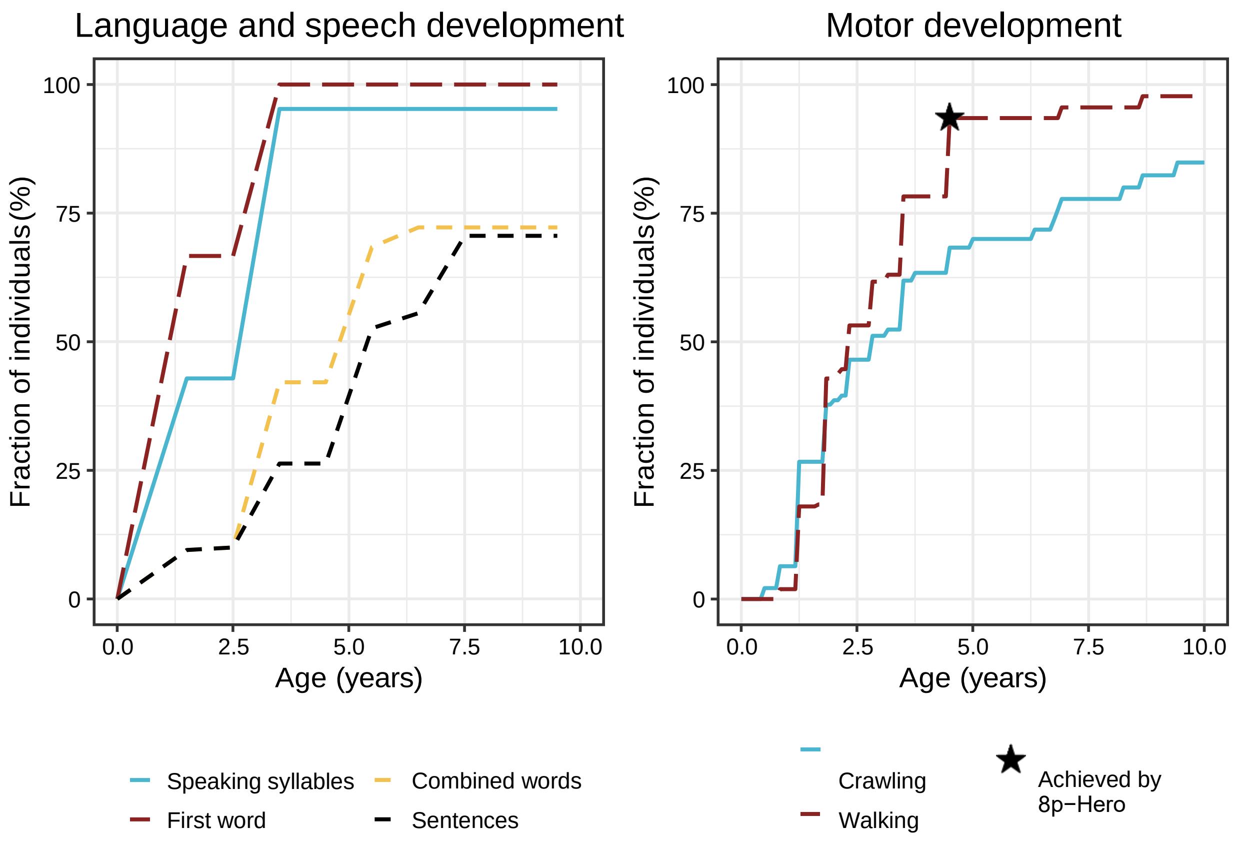


**For developmental milestones achieved, the 8p hero comparison shown is the percentage of 8p heroes that have achieved that milestone by the same age as your 8p hero. If the age when the 8p hero achieved the milestone is known it is indicated by a star.*

### How to interpret symptom frequency plots

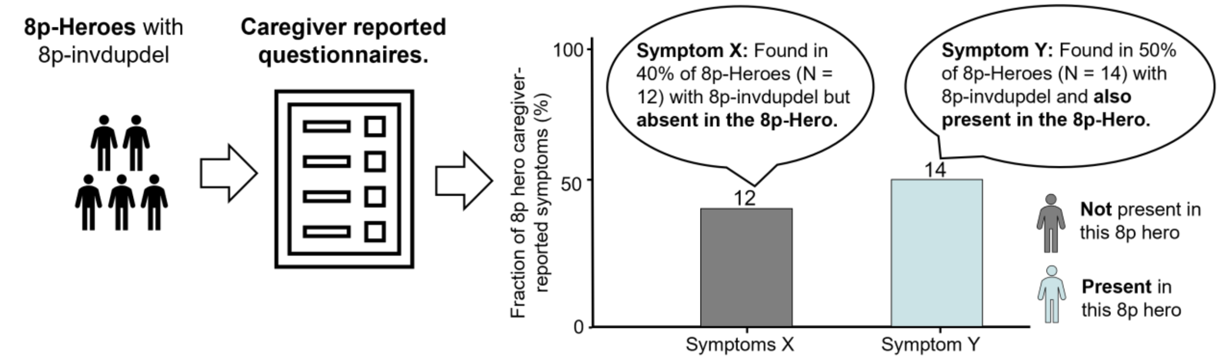


### Physical Health

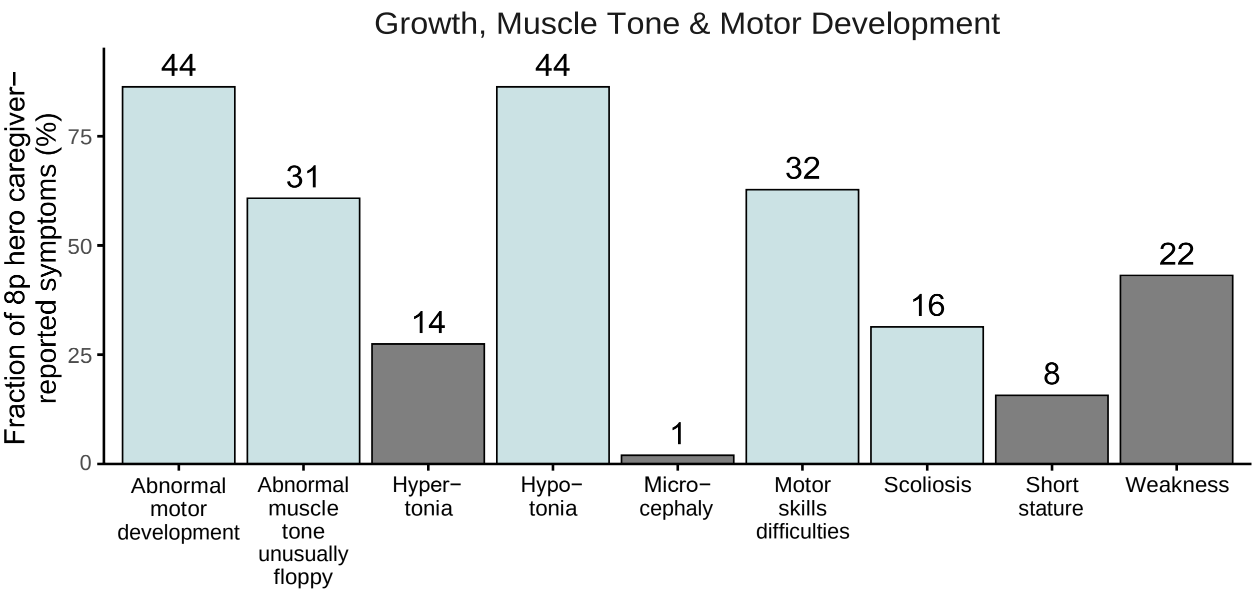


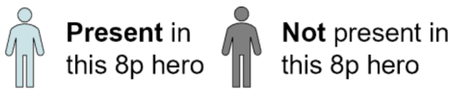

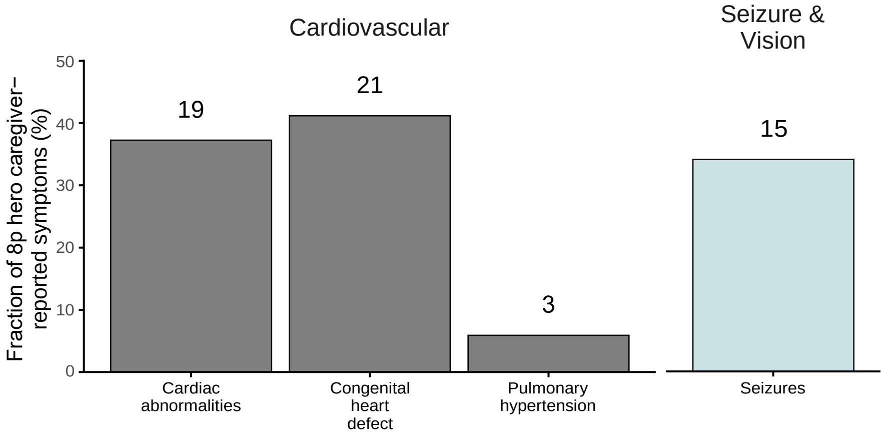


**Present** in this 8p hero **Not** present in this 8p hero

*The age of assessment of the 8p hero was 20-29 years. Please note that the comparison group includes all 8p heroes with 8p inverted duplication/deletions. Be aware that some 8p heroes may not display a clinical characteristic at the time of assessment but might develop it later. The fraction of 8p heroes with a clinical characteristic might thus be underreported. The number above the bar indicates the number of 8p heroes with the clinical characteristic.*

#### Behavioral and Cognitive Development

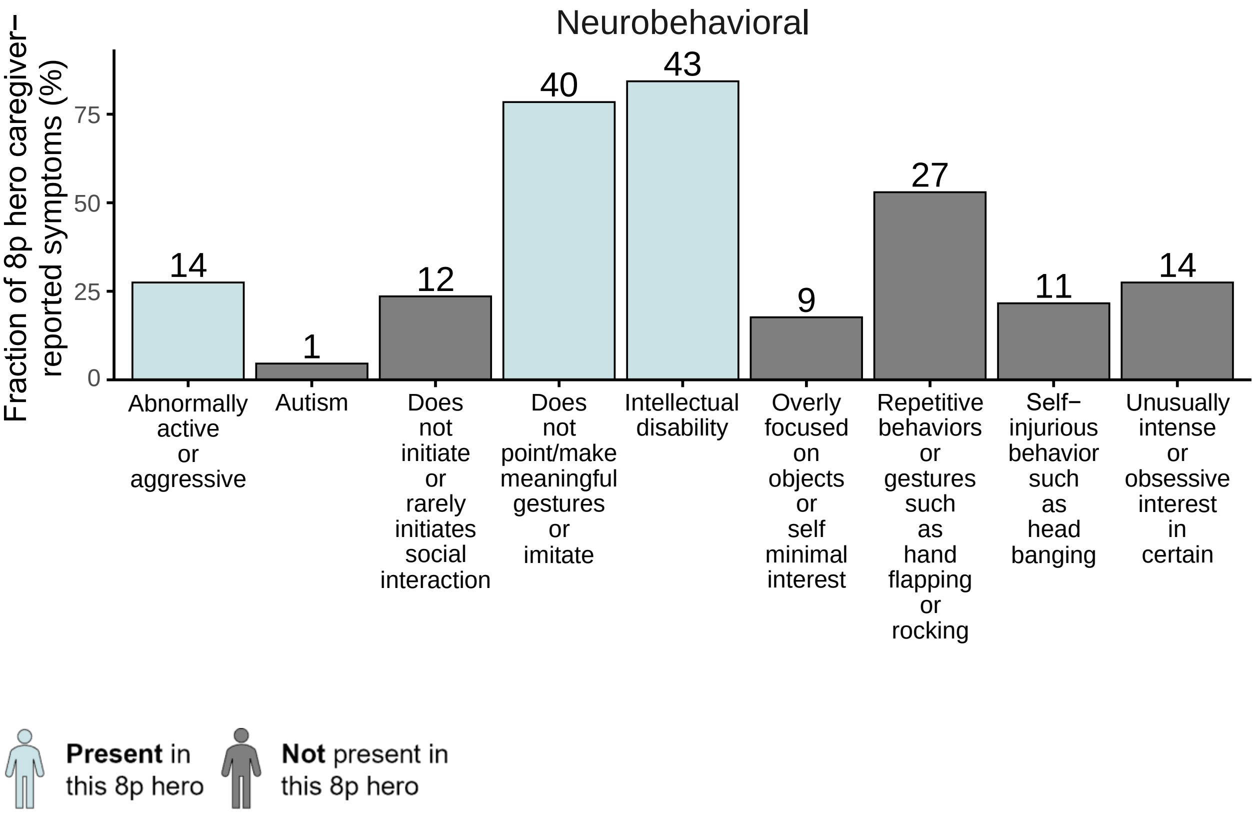


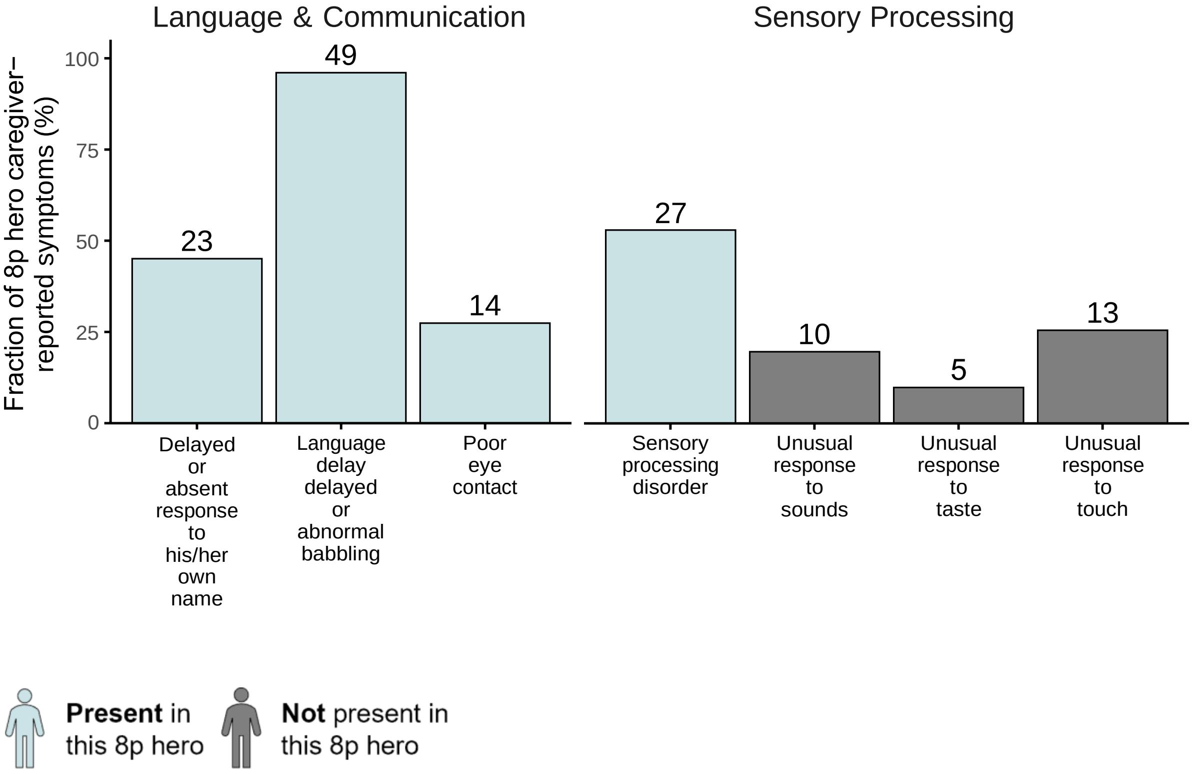


If you have any questions about any of the information provided in this 8p Hero Passport, or would like any of your information updated, please contact us at

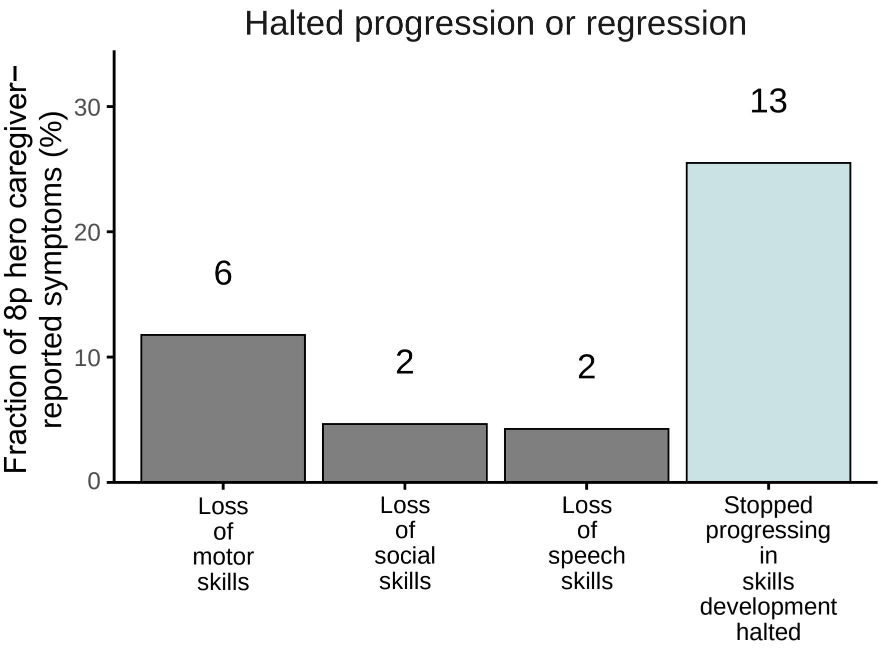


**Present** in this 8p hero **Not** present in this 8p hero

*The age of assessment of the 8p hero was 20-29 years. Please note that the comparison group includes all 8p heroes with 8p inverted duplication/deletions. Be aware that some 8p heroes may not display a clinical characteristic at the time of assessment but might develop it later. The fraction of 8p heroes with a clinical characteristic might thus be underreported. The number above the bar indicates the number of 8p heroes with the clinical characteristic.*

#### Gastrointestinal Health

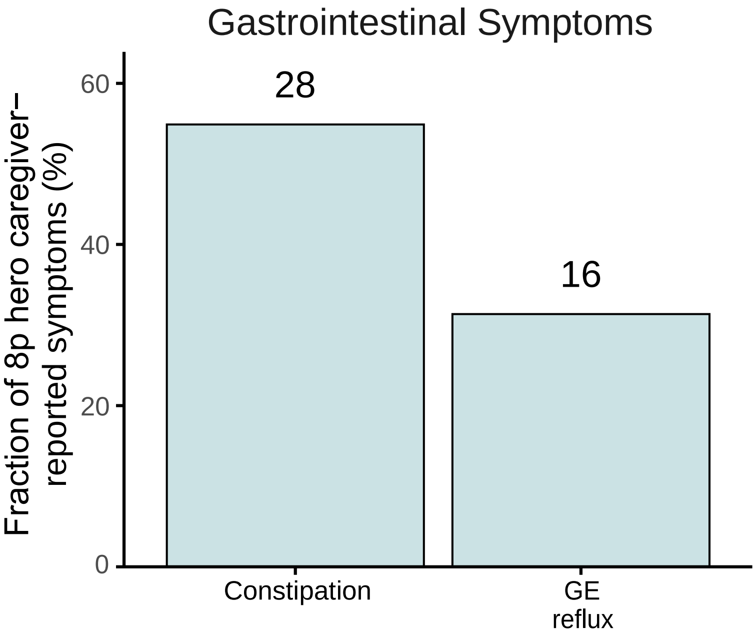


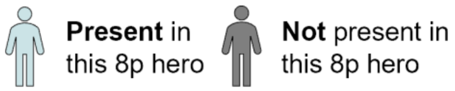


The following Gastrointestinal health symptoms have been documented in the 8p population but we have insufficient data to provide prevalence estimates at this time:

- Diarrhea, jaundice, abdominal pain, enlarged liver, malabsorption, liver function abnormalities, gall-bladder disease, vomiting, cyclic vomiting, inflammatory bowel disease, colic

### Appendix

#### Prior medication and supplements not continued

| Medication | Frequency | Dosage | Stop Reason |
|---|---|---|---|
| Topamax | BID | 1 | NA |
| Pamidronate | treatments | 2 | Finished |
| **Supplement** | **Frequency** | **Dosage** | **Stop Reason** |
| None reported | NA | NA | NA |

### Example 8p Hero Passport: Deletion

**Name:**

**Date of birth:**

**Sex: Female** **Preferred Pronouns: Unknown**

#### Genetic Diagnosis

Date of testing: June-2022 Age of genetic testing: Unknown

Test type: Chromosomal microarray

Genetic finding: Chromosome 8p deletion distal (8p deletion: 8p23.3 (155,000) to 8p23.1 (6,570,000))

Deletion size: 6415000 BP. Median size among other 8p heroes: 4,689,686 BP.

Duplication size: NA

Inheritance: Parental testing not done

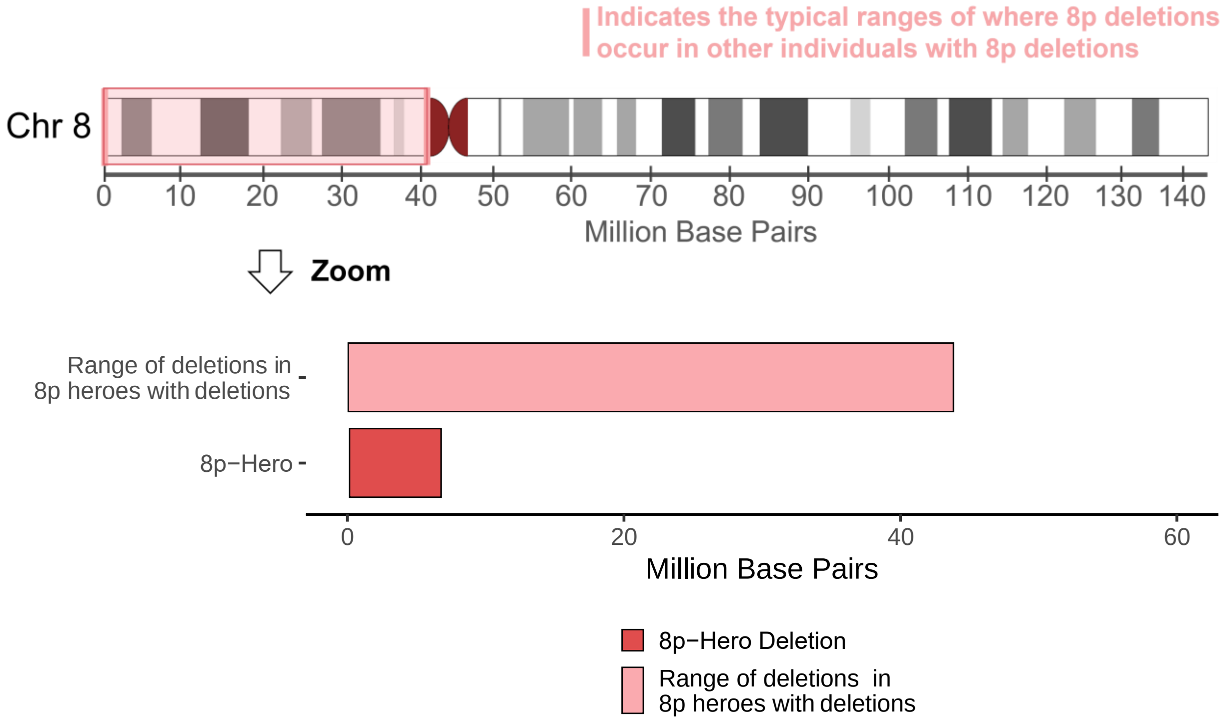


If you have any questions about any of the information provided in this 8p Hero Passport, or would like any of your information updated, please contact us at

#### Family Genetic Testing Recommendation

Given the genetic nature of chromosome 8p disorders, we recommend that family members consider genetic testing to better understand potential hereditary patterns. Family genetic testing can provide valuable insights into the unique genetic makeup of each family member, helping clinicians tailor care more effectively and enabling families to make informed decisions about health and wellness.

For more information on family genetic testing or to discuss options with a genetic counselor, please reach out to.

#### How you can connect with my 8p Hero

When interacting with my 8p Hero, it's important to approach with warmth, patience, and respect. Here are a few ways you can create a meaningful connection:

1. **Start with a greeting:** Say hello clearly and make eye contact when possible. Using my Hero's name helps them recognize your friendly intent.
2. **Pause for a response:** After asking a question or giving an instruction, wait for at least 3 seconds. My Hero may need extra time to process and respond, which could be in the form of a gesture, a nod, or even an eye blink.
3. **Use open body language:** Keep your gestures expressive and welcoming. A warm demeanor often encourages my Hero to feel comfortable in your presence.
4. **Respect personal space but be open to connection:** If my Hero shows interest in a hug or physical touch, depending on your comfort level, it's okay to offer a hand or a gentle hug.
5. **Other ways to connect to my 8p hero:**

#### Chromosome 8p Disorder Summary

Chromosome 8p disorder is a rare genetic condition that is known to affect 550 individuals worldwide, but has a reported prevalence of 1:10,000 - 1:30,000. Chromosome 8p disorder is caused when there is a rearrangement of genetic information before birth, on the short arm (the p arm) of the 8th chromosome. This genetic rearrangement can be the deletion of information, duplication of information, or inverted duplication and deletion (invdupdel). Chromosome 8p disorder has systemic effects, meaning it impacts cells and tissues throughout the body, rather than being confined to a single organ. The chromosomal rearrangement typically arises from spontaneous (de novo) changes early during embryonic development, for reasons that remain unclear. The severity and specific features of 8p disorders can vary significantly based on the amount of genetic information that has changed in addition to other factors that we are still working to understand.

There are several types of therapies that have been found to be effective for children with 8p disorders, depending on their symptoms, these include speech therapy, occupational therapy, physical therapy, vision therapy, ABA therapy, aquatic therapy, feeding therapy, hippotherapy as well as assistive and augmentative communication devices. In the absence of any treatments for chromosome 8p disorders, clinicians at the leading 8p multi-disciplinary clinic recommend treating symptoms with standard clinical practice as they arise.

**Currently, no targeted therapies exist for chromosome 8p disorders. As such, treatment focuses on managing symptoms as they appear, employing standard therapeutic interventions.** If you are treating a patient with chromosome 8p disorder and would like guidance or additional clinical insight from specialists in 8p, please reach out to our team. We can facilitate collaboration with our clinical experts to support your care plan and offer tailored resources. For further assistance, contact our clinical team at.

#### Current Medication & Supplement Information

| Medication | Frequency | Dosage |
|---|---|---|
| None reported | NA | NA |
| **Supplement** | **Frequency** | **Dosage** |
| None reported | NA | NA |

*Prior medications can be obtained from the Appendix.*

#### Prenatal and Birth History

| General information | Your 8p Hero | Other 8p Heroes with 8p deletion median (25-75% quantiles) |
|---|---|---|
| Weight | 2500-4000g (5.5-8.8lbs) | - |
| Length | 45 cm<br>17.7 inch | 49.5cm (46.8cm - 50.8cm)<br>19.5 inch (18.4 inch - 20 inch) |
| Head Circumference | 32 cm<br>12.6 inch | 32.5cm (30.7cm - 33cm)<br>12.8 inch (12.1 inch - 13 inch) |
| Gestational | 37 - 40 weeks | - |
| **Prenatal & birth complications** | | |
| NICU Admission | Yes | 40.7% |
| NICU Reason | Breathing Difficulties | - |
| Days at the Hospital following NICU Admission | 5 | 4 (2 - 8.5) |

#### Developmental Milestones

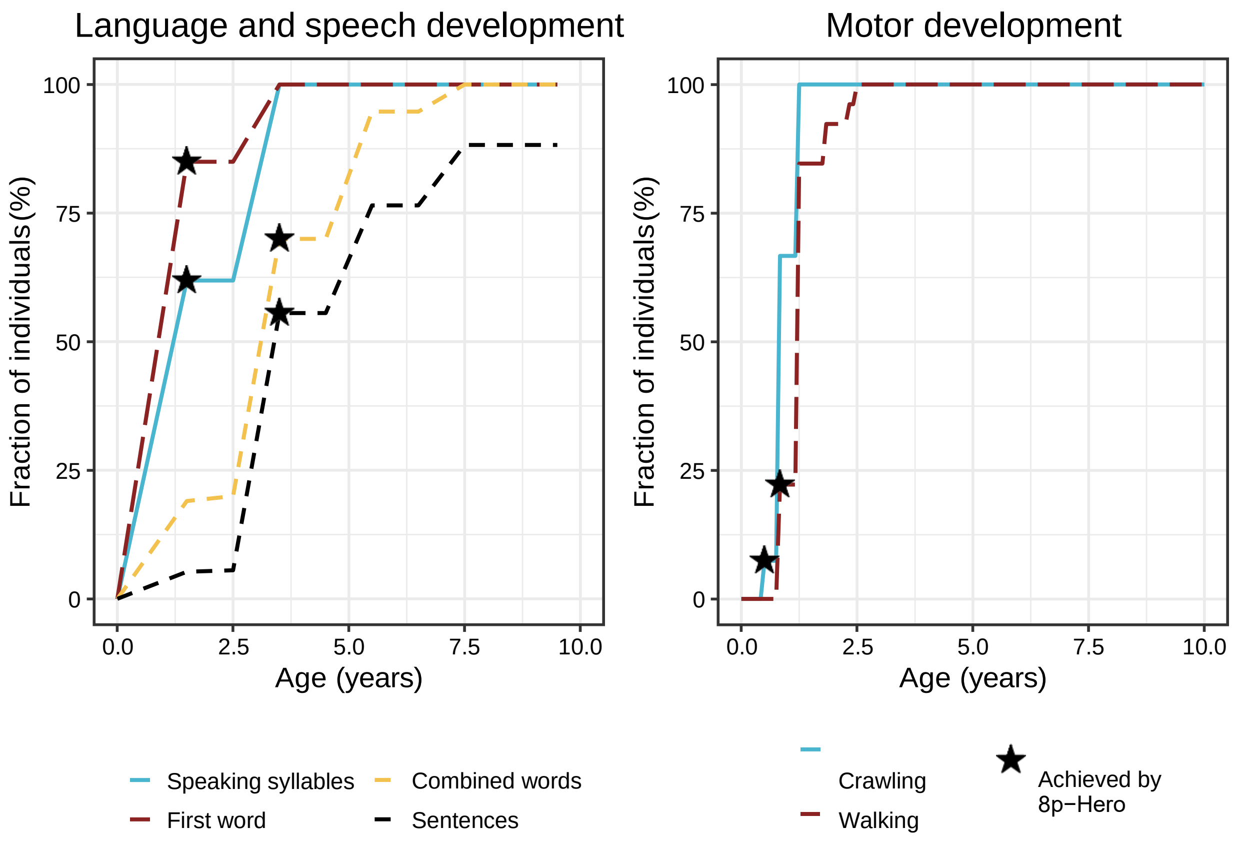


**For developmental milestones achieved, the 8p hero comparison shown is the percentage of 8p heroes that have achieved that milestone by the same age as your 8p hero. If the age when the 8p hero achieved the milestone is known it is indicated by a star.*

### How to interpret symptom frequency plots

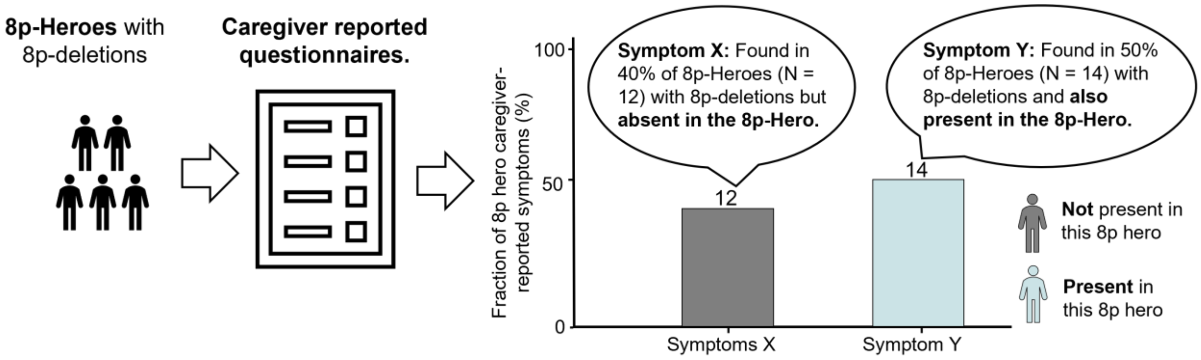


### Physical Health

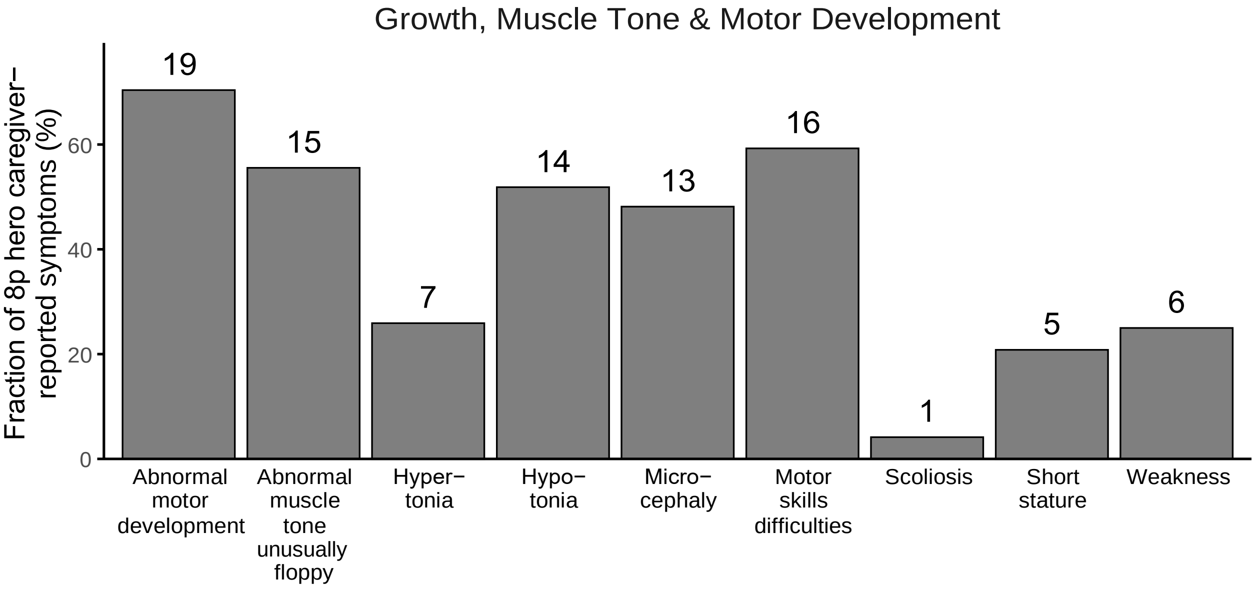


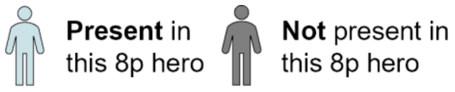


If you have any questions about any of the information provided in this 8p Hero Passport, or would like any of your information updated, please contact us at

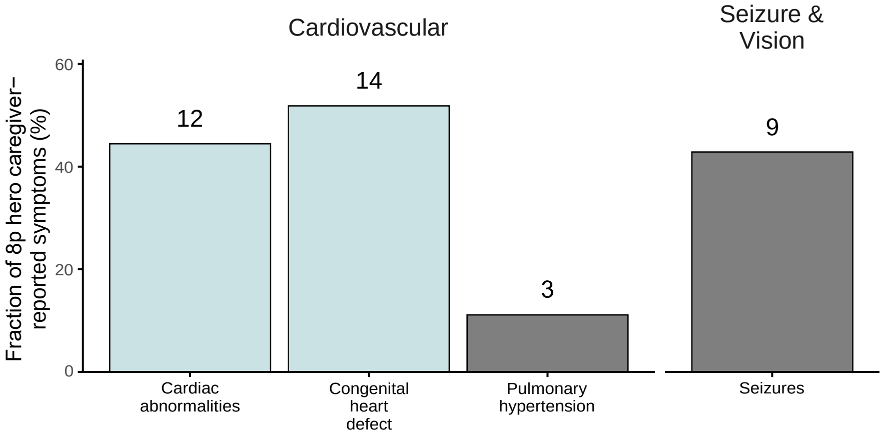


**Present** in this 8p hero

**Not** present in this 8p hero

*The age of assessment of the 8p hero was 20-29 years. Please note that the comparison group includes all 8p heroes with 8p deletions. Be aware that some 8p heroes may not display a clinical characteristic at the time of assessment but might develop it later. The fraction of 8p heroes with a clinical characteristic might thus be underreported. The number above the bar indicates the number of 8p heroes with the clinical characteristic.*

### Behavioral and Cognitive Development

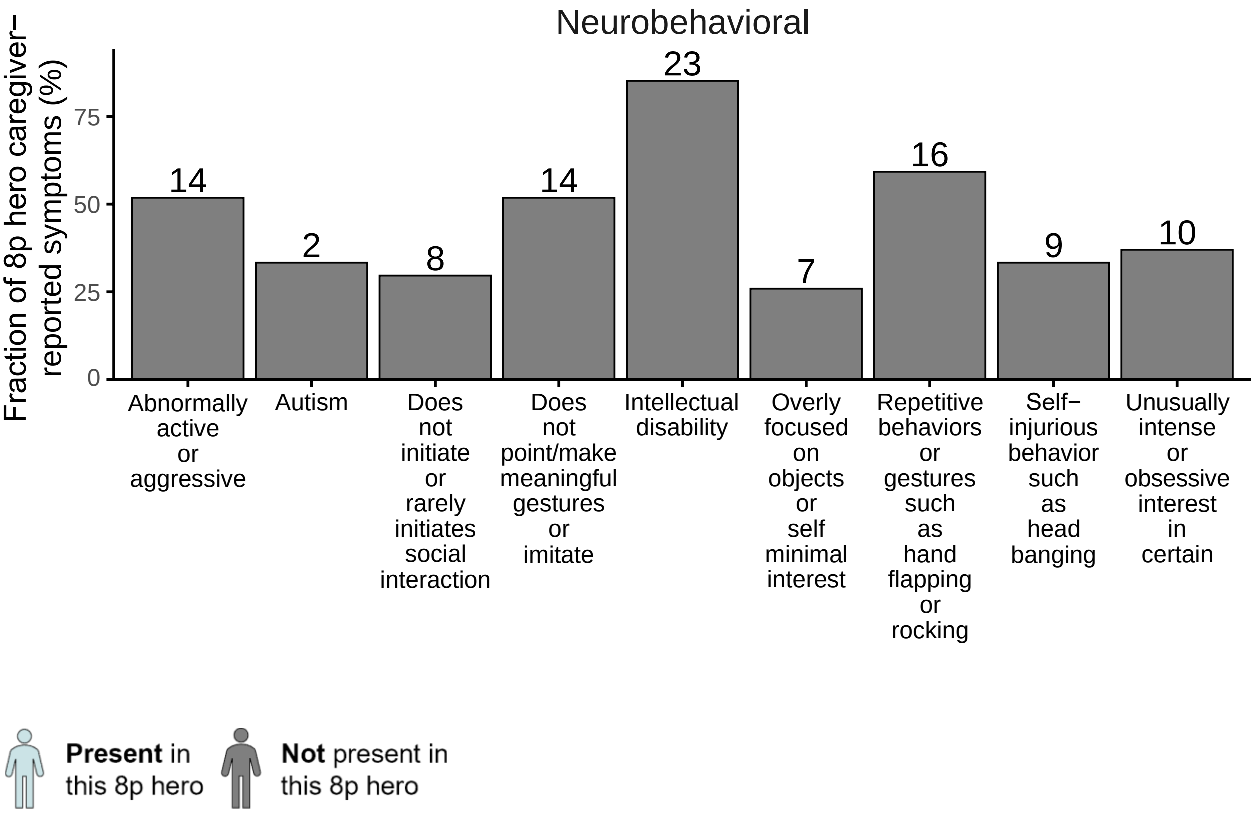


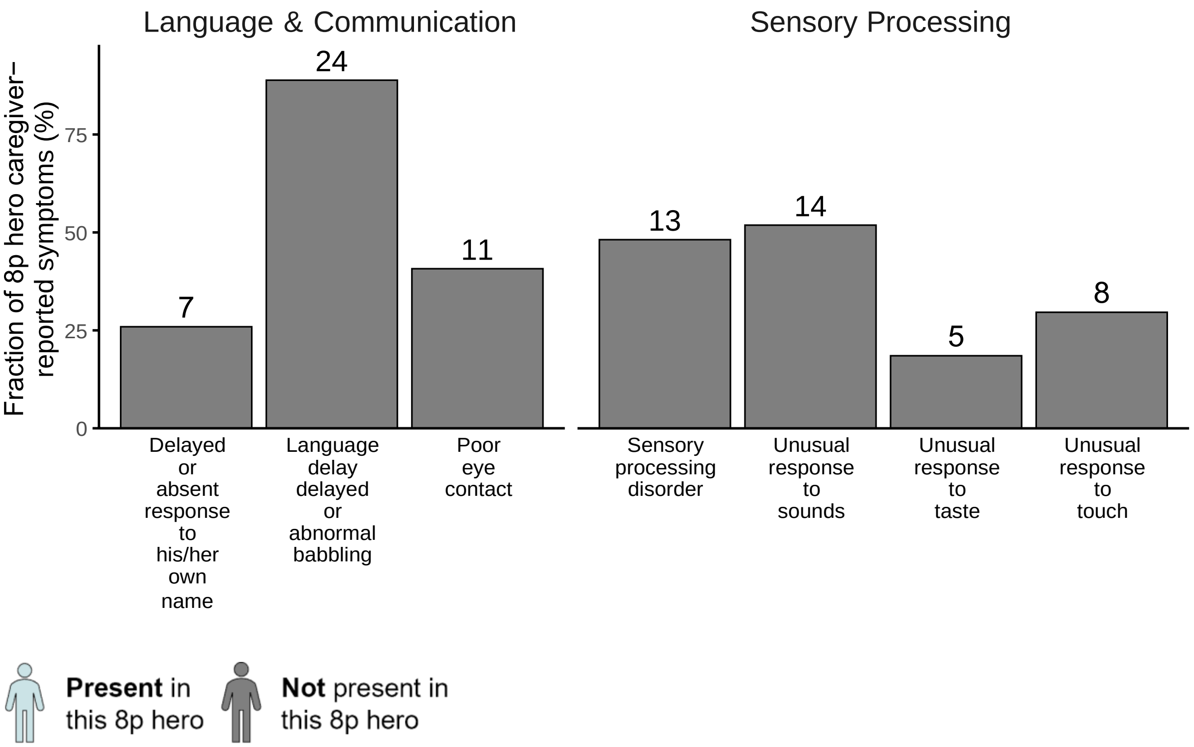


If you have any questions about any of the information provided in this 8p Hero Passport, or would like any of your information updated, please contact us at

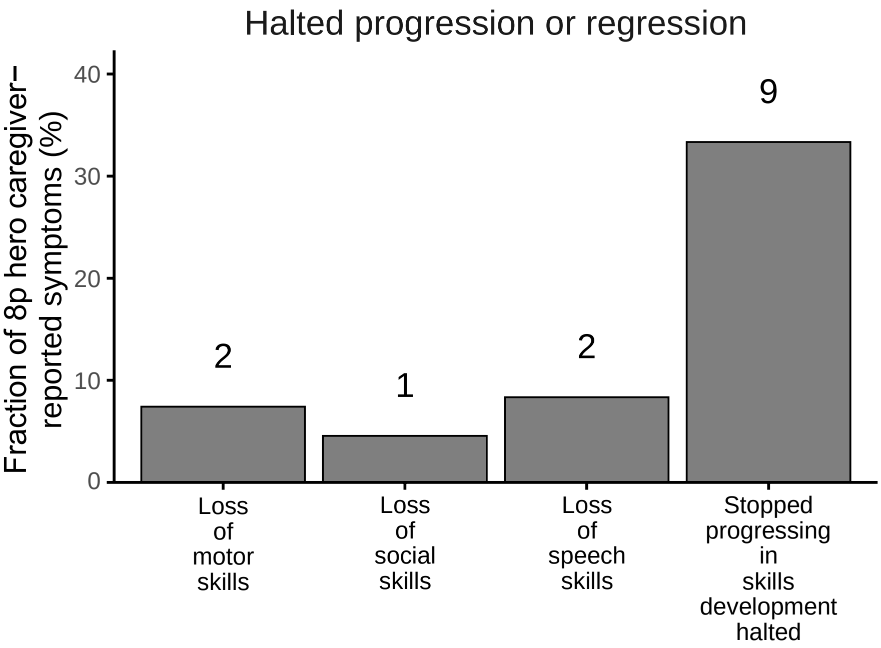


**Present** in this 8p hero **Not** present in this 8p hero

*The age of assessment of the 8p hero was 20-29 years. Please note that the comparison group includes all 8p heroes with 8p deletions. Be aware that some 8p heroes may not display a clinical characteristic at the time of assessment but might develop it later. The fraction of 8p heroes with a clinical characteristic might thus be underreported. The number above the bar indicates the number of 8p heroes with the clinical characteristic.*

#### Gastrointestinal Health

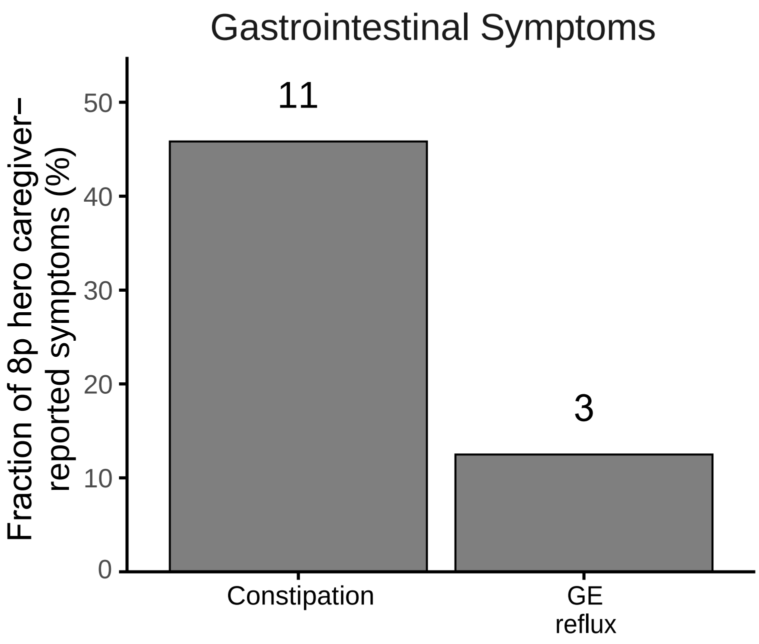


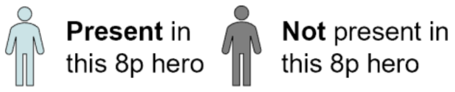


The following Gastrointestinal health symptoms have been documented in the 8p population but we have insufficient data to provide prevalence estimates at this time:

- Diarrhea, jaundice, abdominal pain, enlarged liver, malabsorption, liver function abnormalities, gall-bladder disease, vomiting, cyclic vomiting, inflammatory bowel disease, colic

### Appendix

#### Prior medication and supplements not continued

| Medication | Frequency | Dosage | Stop Reason |
|---|---|---|---|
| None reported | NA | NA | NA |
| **Supplement** | **Frequency** | **Dosage** | **Stop Reason** |
| None reported | NA | NA | NA |

### Example 8p Hero Passport: Duplication

**Name:**

**Date of birth:**

**Sex: Male** **Preferred Pronouns: Unknown**

#### Genetic Diagnosis

Date of testing: Jan-2020 Age of genetic testing: Unknown

Test type: Chromosomal microarray

Genetic finding: Chromosome 8p duplication (Details unknown)

Deletion size: NA

Duplication size: NA

Inheritance: De-novo

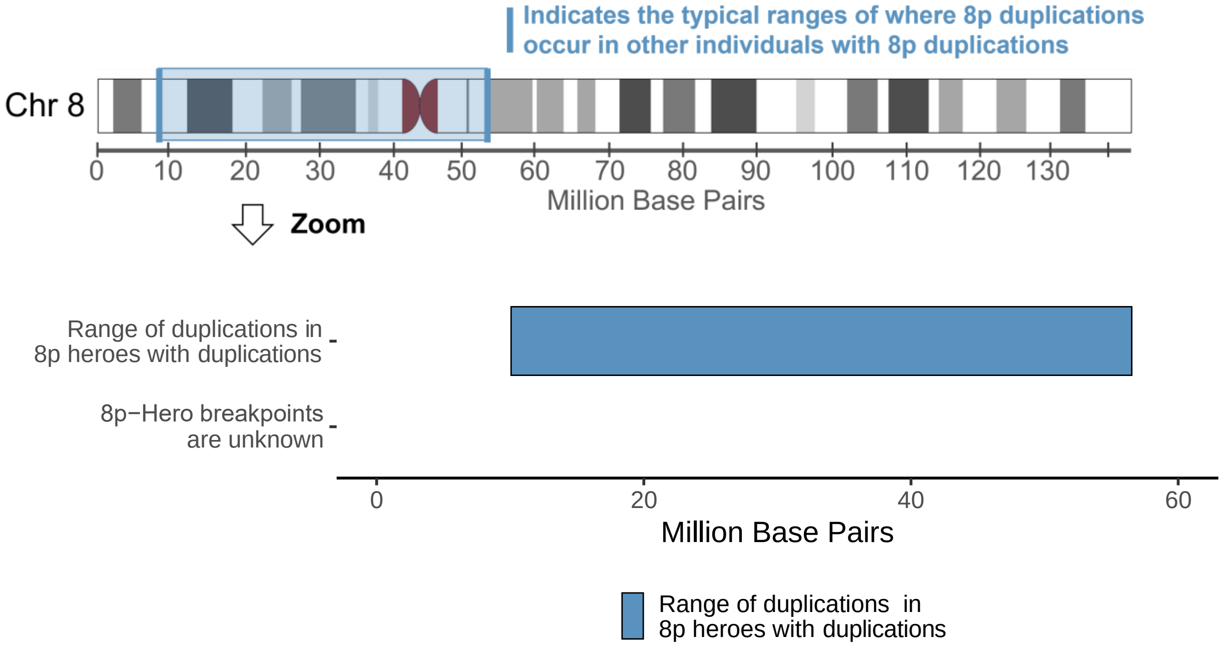


If you have any questions about any of the information provided in this 8p Hero Passport, or would like any of your information updated, please contact us at

#### Family Genetic Testing Recommendation

Given the genetic nature of chromosome 8p disorders, we recommend that family members consider genetic testing to better understand potential hereditary patterns. Family genetic testing can provide valuable insights into the unique genetic makeup of each family member, helping clinicians tailor care more effectively and enabling families to make informed decisions about health and wellness.

For more information on family genetic testing or to discuss options with a genetic counselor, please reach out to.

#### How you can connect with my 8p Hero

When interacting with my 8p Hero, it's important to approach with warmth, patience, and respect. Here are a few ways you can create a meaningful connection:

1. **Start with a greeting:** Say hello clearly and make eye contact when possible. Using my Hero's name helps them recognize your friendly intent.
2. **Pause for a response:** After asking a question or giving an instruction, wait for at least 3 seconds. My Hero may need extra time to process and respond, which could be in the form of a gesture, a nod, or even an eye blink.
3. **Use open body language:** Keep your gestures expressive and welcoming. A warm demeanor often encourages my Hero to feel comfortable in your presence.
4. **Respect personal space but be open to connection:** If my Hero shows interest in a hug or physical touch, depending on your comfort level, it's okay to offer a hand or a gentle hug.
5. **Other ways to connect to my 8p hero:**

#### Chromosome 8p Disorder Summary

Chromosome 8p disorder is a rare genetic condition that is known to affect 550 individuals worldwide, but has a reported prevalence of 1:10,000 - 1:30,000. Chromosome 8p disorder is caused when there is a rearrangement of genetic information before birth, on the short arm (the p arm) of the 8th chromosome. This genetic rearrangement can be the deletion of information, duplication of information, or inverted duplication and deletion (invdupdel). Chromosome 8p disorder has systemic effects, meaning it impacts cells and tissues throughout the body, rather than being confined to a single organ. The chromosomal rearrangement typically arises from spontaneous (de novo) changes early during embryonic development, for reasons that remain unclear. The severity and specific features of 8p disorders can vary significantly based on the amount of genetic information that has changed in addition to other factors that we are still working to understand.

There are several types of therapies that have been found to be effective for children with 8p disorders, depending on their symptoms, these include speech therapy, occupational therapy, physical therapy, vision therapy, ABA therapy, aquatic therapy, feeding therapy, hippotherapy as well as assistive and augmentative communication devices. In the absence of any treatments for chromosome 8p disorders, clinicians at the leading 8p multi-disciplinary clinic recommend treating symptoms with standard clinical practice as they arise.

**Currently, no targeted therapies exist for chromosome 8p disorders. As such, treatment focuses on managing symptoms as they appear, employing standard therapeutic interventions.** If you are treating a patient with chromosome 8p disorder and would like guidance or additional clinical insight from specialists in 8p, please reach out to our team. We can facilitate collaboration with our clinical experts to support your care plan and offer tailored resources. For further assistance, contact our clinical team at.

#### Current Medication & Supplement Information

| **Medication** | **Frequency** | **Dosage** |
|---|---|---|
| None reported | NA | NA |
| **Supplement** | **Frequency** | **Dosage** |
| None reported | NA | NA |

*Prior medications can be obtained from the Appendix.*

#### Prenatal and Birth History

| General information | Your 8p Hero | Other 8p Heroes with 8p duplication median (25-75% quantiles) |
|---|---|---|
| Weight | 2500-4000g (5.5-8.8lbs) | - |
| Length | 45 cm 17.7 inch | 51cm (51cm - 54.6cm) 20.1 inch (20.1 inch - 21.5 inch) |
| Head Circumference | Survey incomplete | 40.8cm (38.4cm - 43.1cm) 16.1 inch (15.1 inch - 17 inch) |
| Gestational | 37 - 40 weeks | - |
| **Prenatal & birth complications** | | |
| NICU Admission | No | 28.6% |
| NICU Reason | Unknown | - |
| Days at the Hospital following NICU Admission | - | 3 (1.5 - 6) |

#### Developmental Milestones

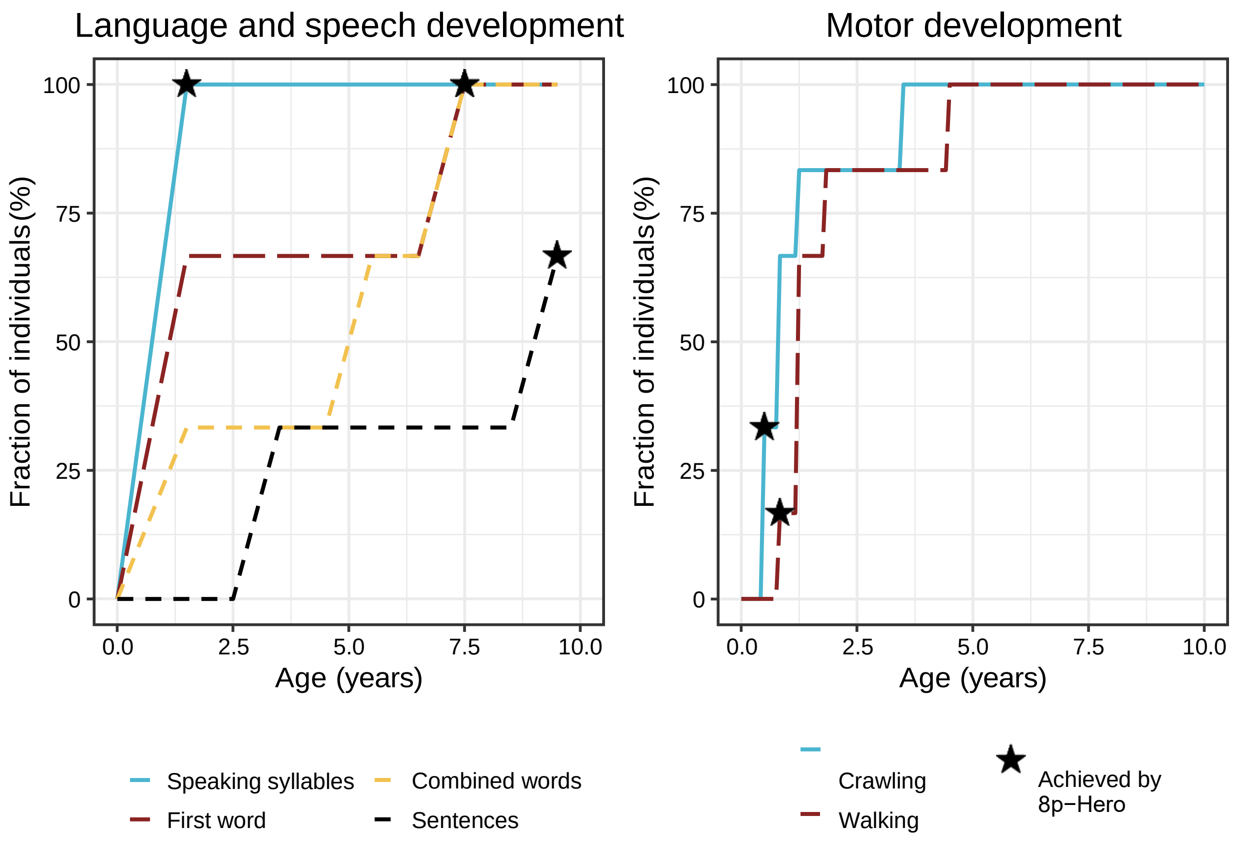


**For developmental milestones achieved, the 8p hero comparison shown is the percentage of 8p heroes that have achieved that milestone by the same age as your 8p hero. If the age when the 8p hero achieved the milestone is known it is indicated by a star.*

If you have any questions about any of the information provided in this 8p Hero Passport, or would like any of your information updated, please contact us at

#### How to interpret symptom frequency plots

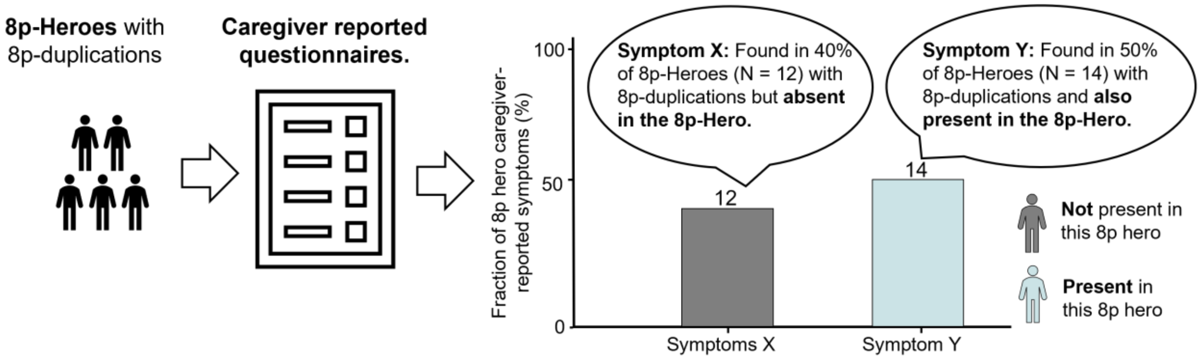


#### Physical Health

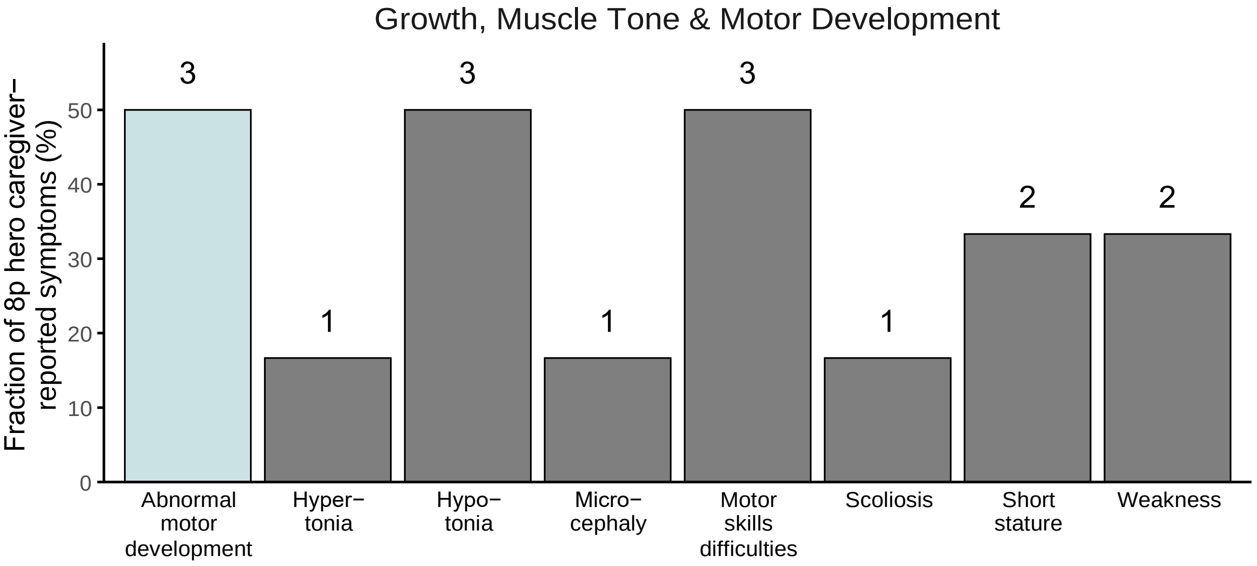


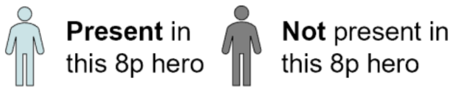

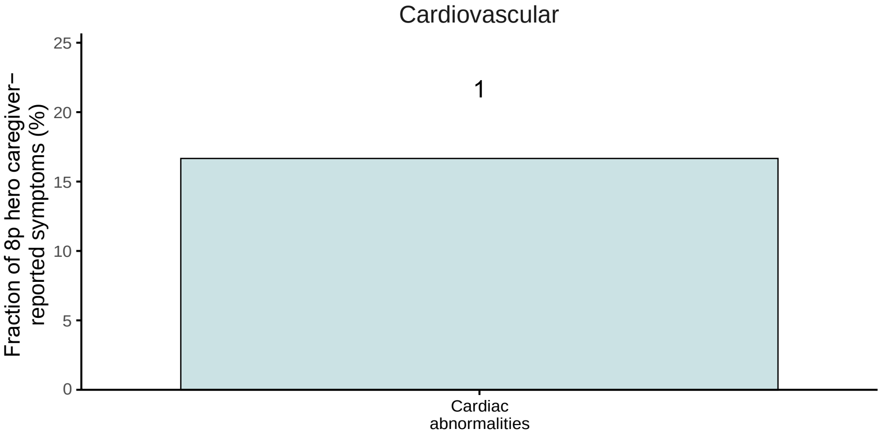


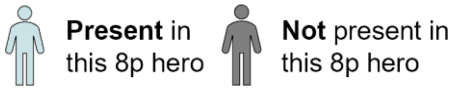


*The age of assessment of the 8p hero was 10-19 years. Please note that the comparison group includes all 8p heroes with 8p duplications. Be aware that some 8p heroes may not display a clinical characteristic at the time of assessment but might develop it later. The fraction of 8p heroes with a clinical characteristic might thus be underreported. The number above the bar indicates the number of 8p heroes with the clinical characteristic.*

### Behavioral and Cognitive Development

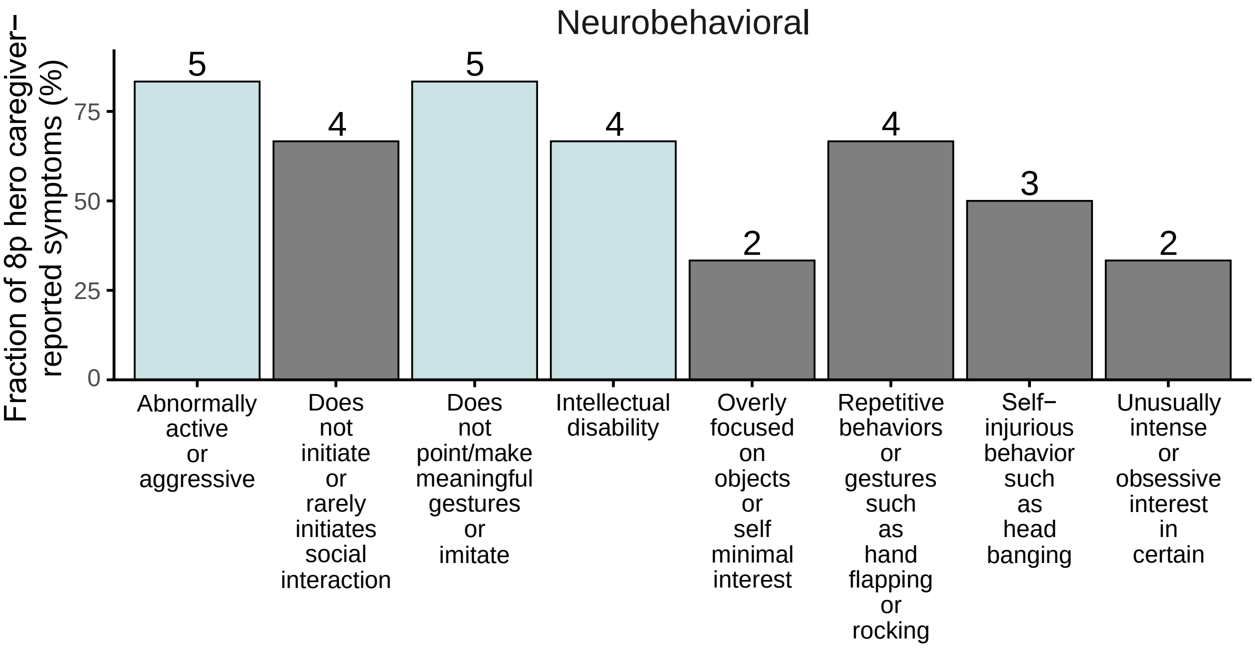


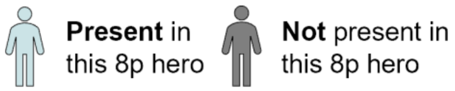


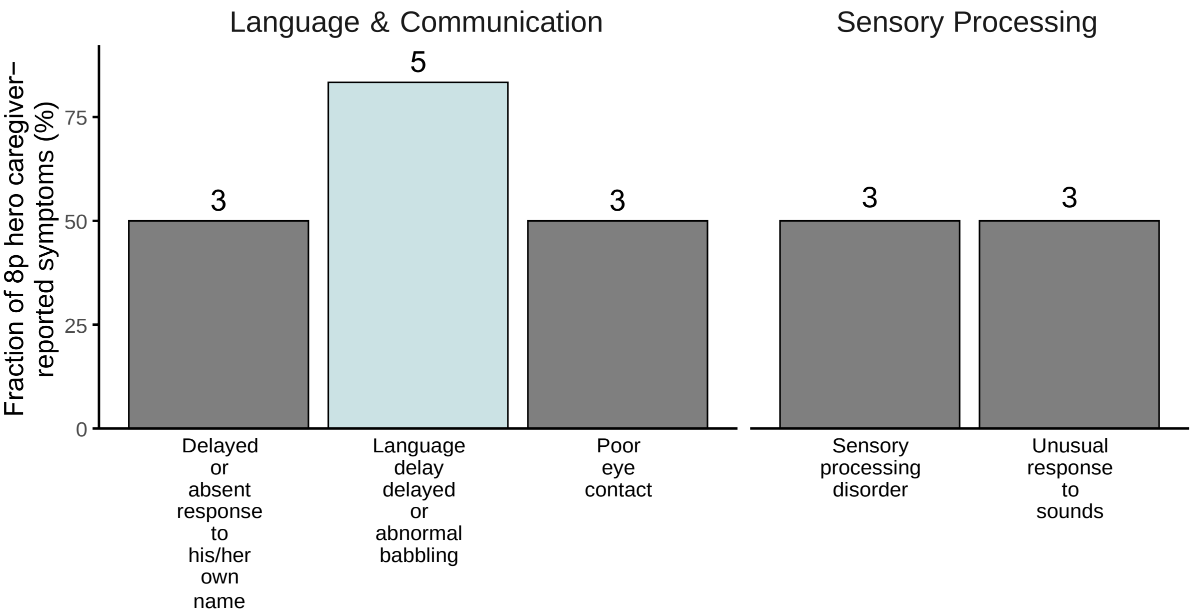


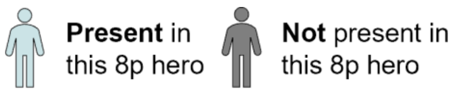


If you have any questions about any of the information provided in this 8p Hero Passport, or would like any of your information updated, please contact us at

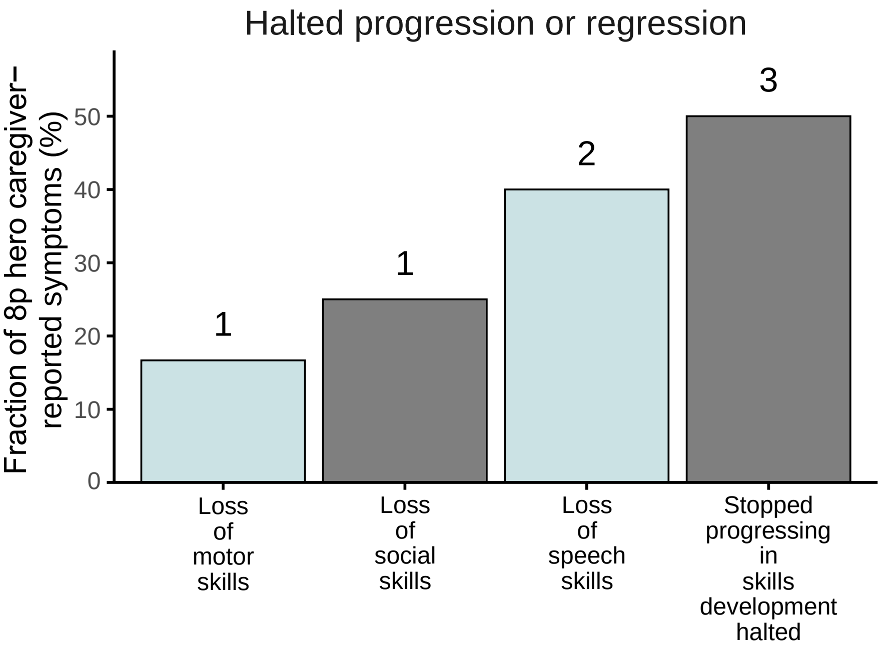


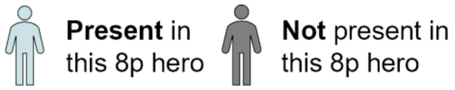


*The age of assessment of the 8p hero was 10-19 years. Please note that the comparison group includes all 8p heroes with 8p duplications. Be aware that some 8p heroes may not display a clinical characteristic at the time of assessment but might develop it later. The fraction of 8p heroes with a clinical characteristic might thus be underreported. The number above the bar indicates the number of 8p heroes with the clinical characteristic.*

#### Gastrointestinal Health

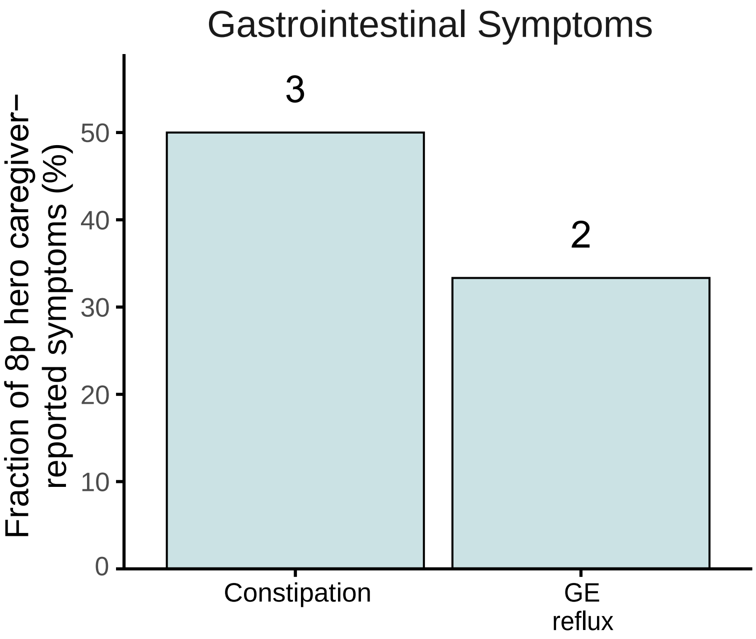


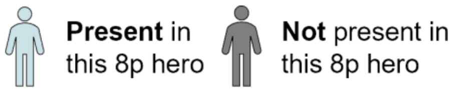


The following Gastrointestinal health symptoms have been documented in the 8p population but we have insufficient data to provide prevalence estimates at this time:

- Diarrhea, jaundice, abdominal pain, enlarged liver, malabsorption, liver function abnormalities, gall-bladder disease, vomiting, cyclic vomiting, inflammatory bowel disease, colic

### Appendix

#### Prior medication and supplements not continued

| Medication | Frequency | Dosage | Stop Reason |
| --- | --- | --- | --- |
| None reported | NA | NA | NA |
| **Supplement** | **Frequency** | **Dosage** | **Stop Reason** |
| None reported | NA | NA | NA |
